# Contact network structure shaped pandemic transmission despite lockdowns

**DOI:** 10.64898/2026.02.06.26345745

**Authors:** Zhi Ling, Shozen Dan, Yu Chen, Veronika Jaeger, André Karch, Oliver Ratmann, Swapnil Mishra

## Abstract

Human contact network structure fundamentally shapes infectious disease transmission and control. Most COVID-19 epidemic models assumed approximately homogeneous contact patterns, yet real-world networks are highly heterogeneous. We analysed 59,585 daily non-household contact reports from Germany’s COVIMOD study (2020–2021) using a novel heavy-tail regression framework. Throughout the pandemic, contact distributions remained strongly heavy-tailed despite substantial non-pharmaceutical interventions. The most connected 20% of individuals consistently generated over 80% of non-household contacts, placing the network in an infinite-variance, scale-free regime for all but the first lockdown. Regions with heavier tails experienced higher COVID-19 incidence. Importantly, the top 5% quantile had just 9.4 contacts per day, reflecting everyday activities rather than mass gatherings. High-contact individuals were predominantly working-age adults living alone and engaged in in-person work or study. Agent-based simulations show that tail-targeting interventions could control spread as effectively as lockdowns for a broad range of pathogens with far less disruption, with childhood-driven infectious disease remaining an exception. These findings demonstrate that the structure of human contact networks is fundamental to how rapidly infectious diseases spread, persist, and how transmission can be controlled.

---

Epidemics unfold on networks of human contacts, from households and workplaces to long-distance travel, and this structure is central to how pathogens such as mpox, influenze, Ebola and SARS-CoV-2 spread through populations. ^1,2^ Over recent decades, global and local connectivity have intensified through urbanisation, commuting and mobility, creating denser and more complex interaction networks. When a novel pathogen emerges, cutting contacts is therefore one of the first control measures, implemented through stay-at-home orders, limits on gatherings, school and workplace closures and travel restrictions. ^3–5^ Yet it remains unclear how effective such non-pharmaceutical interventions (NPIs) were at fragmenting the underlying contact network: did lockdowns truly dismantle giant connected components of social interaction, or did they mainly thin out already sparse links while highly connected individuals remained active?

Human social networks are far from random. Their topology shapes not only how fast an epidemic starts but also its eventual size and the conditions under which it dies out. A key organising principle is the distribution of the number of social contacts per person per day (the degree distribution). This distribution is typically highly right-skewed, with most individuals having few contacts and a minority acting as hubs that may sustain superspreading through their contacts. ^1,6^ When the degree distribution follows a power law *P* (*k*) ∝ *k*^−ω^ for large degree *k*, the value of the tail exponent *ω* determines the moment structure. For *ω >* 3, the variance of degree is finite, corresponding to a relatively homogeneous, finite-variance regime; for 2 *< ω <* 3, the variance diverges in the infinite-population limit, placing the network in an infinite-variance regime in which a small number of hubs dominate connectivity and fundamentally alter transmission dynamics compared to standard SIR-type compartmental models. In particular, on infinite-variance networks, epidemics can spread rapidly and widely even when the average number of contacts is low, as the reproduction number directly scales with the second moment of the degree distribution in very large populations and the epidemic threshold vanishes. ^7–9^

This same non-random structure that accelerates spread also creates opportunities for interventions to disrupt transmission. Targeting hubs or highly connected settings can in principle curb transmission more efficiently than uniform reductions in contact, and may allow less socially and economically disruptive control strategies than population-wide lockdowns. ^7,9^ During COVID-19, a range of such strategies were proposed or implemented, including circuit-breaker interventions, household bubbles and pods, hybrid working and workplace flexibility, digital exposure notification apps, and refined contact tracing supported by mobile technologies. ^10,11^ The effectiveness of these approaches ultimately depends on the evolving structure of contact networks: who remains highly connected under different NPIs, how the tail of the contact distribution responds, and whether interventions move the network from an infinite-variance to a finite-variance regime.

Several data sources provide partial views of these networks. High-resolution proximity data from wearable sensors or WiFi logs on campuses, workplaces and hospital wards reveal local patterns of face-to-face encounters. ^12–14^ Large mobility and social media datasets have been used to infer longer-range connections and changes in movement patterns during the pandemic; ^15,16^ however, the mapping from such signals to actual face-to-face contacts relevant for respiratory pathogen transmission remains uncertain. ^17^ During the COVID-19 pandemic, Europe and other regions implemented repeated, population-representative social contact surveys that directly asked participants to report their recent contacts. ^18–22^ A key innovation in these surveys is the ability to record group contacts, addressing at source the difficulty respondents face in listing every single person they meet. To date, analyses of these rich datasets have predominantly focused on average contact rates and age-specific mixing patterns. ^19,22^

Here, we characterise the full shape of the social contact distribution throughout the COVID-19 pandemic across Germany, and its tail behaviour coupled to network properties and epidemic transmission dynamics. We focus on non-household contacts, defined as face-to-face conversations or physical encounters outside the respondent’s own household. This choice reflects both epidemiological and practical considerations: non-household contacts are the main target of NPIs such as workplace closures and restrictions on social gatherings, whereas household contacts are difficult to change without substantial social cost. Methodologically, we introduce a distributional regression framework that builds on the Beta–Negative Binomial (BNB) likelihood. ^10^ The BNB generalises the Negative Binomial and Waring families and spans Poisson-like, thin-tailed behaviour to heavy-tailed power-law-like behaviour. To facilitate modelling, we express the BNB directly through the mean, overdispersion and a tail index analogous to the tail exponent *ω*. This allows us to infer whether and when the contact network lies in a finite-or infinite-variance regime, while naturally accommodating deviations from a pure power law at low degrees. We embed the BNB likelihood in a semi-parametric Bayesian multilevel model with age-, sex- and time-varying effects, and we use post-stratification to recover population-level degree distributions and their uncertainty. Leveraging individual-level, large-scale, and longitudinal COVIMOD contact survey data over a period that encompassed multiple epidemic waves of SARS-CoV-2 and a range of NPIs, ^22,24^ we address three questions. First, did non-household contact distributions in Germany during COVID-19 occupy finite-or infinite-variance regimes at different phases of the pandemic, and how persistently heavy-tailed were they? Second, which demographic, behavioural and attitudinal factors are associated with membership in the high-contact tail of the distribution? Third, how do these empirically grounded degree distributions affect epidemic outcomes under different intervention strategies? To answer the latter, we simulate the spread of template pathogens, including SARS-CoV-2 ancestral and Omicron variants and pandemic influenza A (H3N2), on synthetic networks calibrated to the inferred degree distributions. Together, these analyses link the evolving heavy tails of human contact patterns to both the vulnerabilities and the opportunities for more precise, less disruptive epidemic control.

## Results

### Social contact distributions remained heavy-tailed

During 2020–2021, Germany went through repeated phases in which daily case counts grew, stabilised or declined (Fig. 1a), primarily reflecting the success of non-pharmaceutical interventions (NPIs) in reducing average transmission. The COVIMOD study is a nationwide contact survey covering all NUTS 3 (nomenclature of territorial units for statistics, level 3) regions in Germany (Fig. 1b), recruited based on age, sex, and region quotas to ensure sample representativeness of the German population. From 30 April 2020 to 31 December 2021, the survey collected 59,585 responses from 7,851 participants. Participants were asked to report social contact information for the past 24 hours, with contact defined as either face-to-face conversation or physical contact, such as handshakes or hugs. ^18,22^ The 33 survey waves can be structured into 5 phases ^24^ comprising the first lockdown, a period of relaxed NPI measures, a second lockdown, another period of relaxation, and a post-vaccination period (Fig. 1a).

**Fig. 1.**
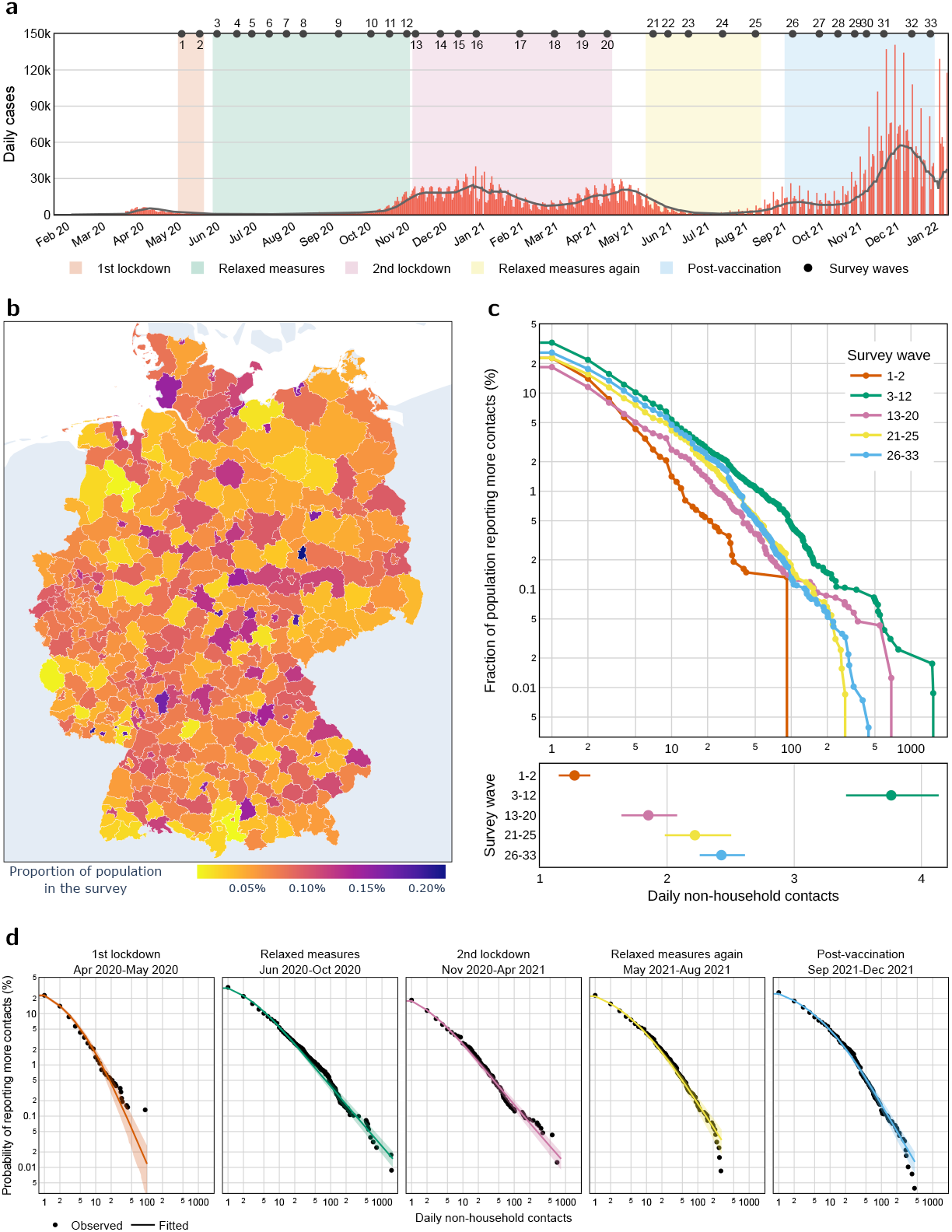
Measures of shifting social contact networks during the COVID-19 pandemic in Germany, measured through the COVIMOD survey. **a**, The COVIMOD survey comprised 33 waves, divided into 5 groups according to the German national-level NPIs. We display the midpoint date of each wave (black solid dots), the daily number of confirmed COVID-19 cases (red bars), and the 7-day rolling average (black line). b, Geographic distribution of survey participation, shown as the proportion of responses (including repeat participation) relative to the population of each NUTS 3 region. c, Upper: observed CCDFs (solid dots) of daily non-household contacts on a log-log scale. Lower: average daily non-household contacts (solid dots: posterior medians, line ranges: 95% CrIs) for each period. d, Comparison of empirical CCDFs (black dots) with the fitted CCDFs (solid lines: posterior medians, shaded areas: 95% CrIs). The dashed line indicates a reference scale-free regime and slope. In panels c, and d, color codes follow that of panel a. In panels c and d, observations, estimates and intervals are post-stratified to match the age-sex distribution of the German population.

Across phases, we observed striking heterogeneity in the number of daily non-household contacts (Fig. 1c). Prior to COVID-19, the average number of non-household contacts ranged between 6-16 among adults, ^18^ and these dropped to 1-4 across all phases. To investigate the distribution of these contacts and highlight the strength of its heavy tail, we visualized the complementary cumulative distribution function (CCDF) on log–log axes. Even during phases with declining incidence, the right-hand tail decayed slowly and approximately linearly, characteristic of heavy tailed behaviour. In the relaxed summer 2020 period, the top 5% of individuals reported more than 11 non-household contacts in a day, despite an average of only 3.8 contacts and around half of people reporting no non-household contacts at all. In contrast, before the pandemic, the top 5% had more than 16-43 non-household contacts per day across European countries. ^18^ Thus, both the mean and the heavy tail shifted to lower values relative to pre-pandemic levels, and this remained so also in the post-vaccination period.

The near-linear tail of the CCDF suggests an underlying power-law regime. When the tail exponent *ω* lies between 2 and 3, such distributions are in an infinite-variance regime, implying that a small fraction of hubs dominates connectivity. To quantify this structure over time, we fitted a Bayesian multilevel “heavy-tail regression” model in which individual contact counts follow a Beta–Negative Binomial (BNB) distribution. We inferred age- and sex-specific degree distributions for each phase, using Gaussian process priors over age and post-stratification to the German population (Methods). In leave-one-out cross-validation, BNB-based models outperformed classic discrete power-law models and thin-tailed alternatives (Supplementary Fig. 1 and Supplementary Table 1), and thin-tailed candidates also failed posterior predictive model consistency checks. ^26^

Figure 1d demonstrates that this approach accurately captures the national contact distribution across its full range, from zero to over 400 non-household contacts per day, across all five, highly heterogeneous phases. Germany’s non-household contact distribution remained heavy-tailed throughout the study period. High-degree individuals consistently followed a power law, whereas low-degree individuals occurred less frequently than a power law would predict—a pattern that has been theoretically explained, ^26^ observed in previous studies, ^27,28^ and correctly reproduced by our model. We found that the transition from the non-power-law body to the scale-free tail was gradual, highlighting limitations in conventional cutoff-type or exceedances fitting approaches. ^29^ Nevertheless, an approximate *k*_min_ can still be obtained for each period through qualitative assessment (Supplementary Fig. 2), highlighting that a minority of highly connected individuals sustained an infinite-variance tail even during periods when *R*_*t*_ ≲ 1, preserving the structural capacity for superspreading.

### Most contacts persistently originated from twenty percent of the population

We next quantified how much of the population-level mixing burden was carried by the most connected individuals. For each phase, we ordered the respondents by degree and computed the cumulative number of contacts generated by the top *x*% of individuals (for *x* from 0 to 100) and the corresponding share of all contacts (Fig. 2a).

**Fig. 2.**
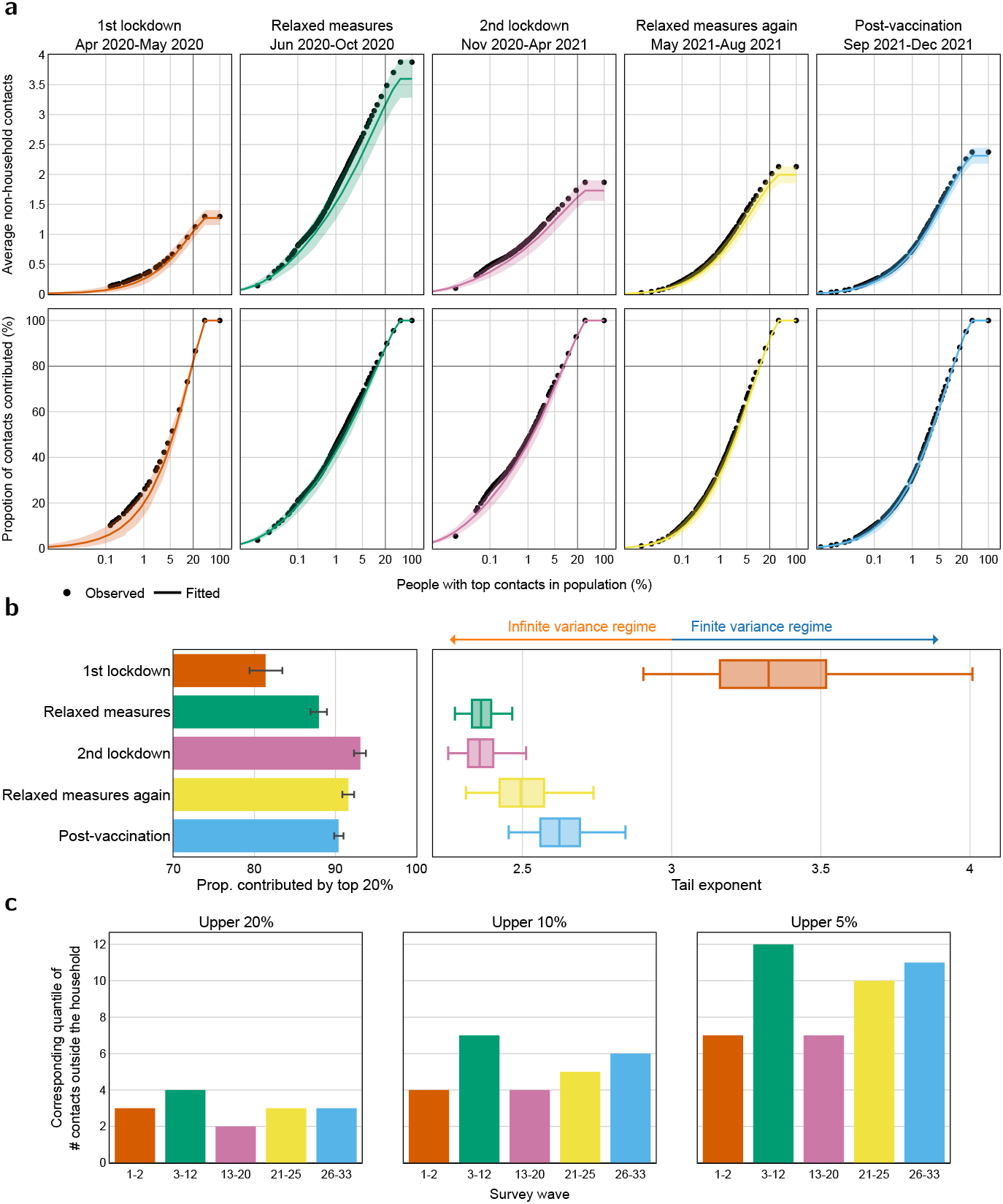
Heavy tail properties, persisting Pareto law, and infinite variance in the degree distribution. **a**, Upper: the average number of non-household contacts contributed by the top *x*% of individuals, ranked in descending order by their number of non-household contacts. Lower: the proportion of non-household contacts contributed by the top *x*% of the population (inverted Lorenz curve). The observed values (black dots) are compared with posterior medians (solid lines) with 95% CrIs (shaded bands). The *x*-axis is on a logarithmic scale. b, Left: the posterior medians (bars) and 95% CrIs (error bars) for the proportion of non-household contacts contributed by the top 20% of the population. Right: estimated tail exponent of the German contact network (bootstrapped median, IQR, and 95% CI). c, Estimated upper 20%, 10%, and 5% percentiles (bars) of the non-household contact distribution. In all three panels, color codes follow that of Fig. 1a. In panels a and b, observations, estimates and intervals are post-stratified to match the age-sex distribution of the German population.

During the first lockdown, the top 20% of most connected individuals accounted for 81.3% of all non-household contacts (posterior median, 95% credible interval 79.3–83.1%). When restrictions were relaxed, this Pareto pattern intensified: the top 20% group generated 87.8% of all contacts. Across the full study period, the top quantile of the population invariably accounted for more than 80% of all non-household contacts (Fig. 2b, left). In other words, the Pareto principle—roughly “80% of contacts come from 20% of people”—held robustly across very different epidemiological and policy contexts.

We then examined how NPIs reshaped the tail of the degree distribution. The key parameter is the estimated tail exponent *ω*. During the first lockdown, we inferred *ω* ≈ 3.32 (95% credible interval 2.90–4.01), placing the network into a finite-variance (random-like) regime (Fig. 2b, right). In this phase, strict population-wide restrictions not only reduced average contacts but also thinned out high-degree nodes sufficiently that the residual network corresponded to a random finite-variance network, greatly supporting the disruption of disease spread. ^7^ As society reopened in June–September 2020, the exponent dropped to *ω* ≈ 2.36 (95% CrI 2.27–2.47), firmly back in the infinite-variance regime, and remained below 3 thereafter. The later second lockdown did not succeed in mitigating the heaviness of the tail as the first lockdown had done. This pattern reveals a form of structural plasticity: NPIs can temporarily push the contact network across a phase boundary between finite- and infinite-variance regimes, but once restrictions ease, the network rapidly self-organises back into a structure on which even weakly transmissible pathogens can spread and epidemics are highly likely to persist. ^1,30^

### Population-level features of heavy tails could enable nuanced tailoring

Next, we evaluated who, demographically and behaviourally, occupied the high-contact tail that drives the Pareto patterns. We did this by identifying the minimum number of contacts corresponding to the top 20%, 10% and 5% of the distribution. Critically, even the top 5% quantile corresponded to just 9.4 non-household contacts per day across all phases, indicating that unlike time periods outside the pandemic, ^31^ during the pandemic and the post-vaccination period the heavy tail reflected everyday activities rather than just individuals involved in mass gatherings or with special occupations, such as teachers or healthcare workers. This prompted us to delineate the group-level characteristics of the individuals in the top 20%, 10% and 5% tail groups (Fig. 2c), as measured in the survey. Fitting a logistic regression with sparsity-inducing priors (Methods), we estimated each factor’s relative risk (RR) of a person being in a high-contact group, compared to the average population risk (Table 1).

**Table 1.** Demographic and behavioural characteristics of being among the most socially connected individuals during the COVID-19 pandemic in Germany.

| Variable | Category | RR 20%<sup>1</sup> | RR 10% | RR 5% |
| --- | --- | --- | --- | --- |
| Household size | 1 person | **1.20** (1.14, 1.25) | 1.06 (0.99, 1.13) | 1.02 (0.94, 1.10) |
|  | 2 person | **1.06** (1.01, 1.10) | 1.04 (0.98, 1.10) | 1.05 (0.97, 1.14) |
|  | 3 person | **0.94** (0.89, 0.98) | 0.96 (0.91, 1.02) | 1.00 (0.93, 1.06) |
|  | 4 person | 0.98 (0.92, 1.02) | 1.00 (0.94, 1.07) | 1.03 (0.96, 1.14) |
|  | 5+ person | **0.86** (0.79, 0.93) | 0.94 (0.85, 1.02) | 0.90 (0.80, 1.02) |
| Employment | Employed, full-time | **1.37** (1.30, 1.44) | **1.75** (1.61, 1.88) | **2.33** (2.06, 2.66) |
|  | Employed, part-time | **1.42** (1.34, 1.51) | **1.71** (1.57, 1.87) | **2.18** (1.88, 2.51) |
|  | Full-time parent | **0.55** (0.46, 0.65) | **0.34** (0.23, 0.45) | **0.20** (0.09, 0.34) |
|  | Long-term sick/disabled | 0.99 (0.87, 1.08) | 0.88 (0.71, 1.05) | 0.92 (0.64, 1.11) |
|  | Retired | 0.97 (0.90, 1.02) | **0.85** (0.76, 0.95) | **0.66** (0.55, 0.79) |
|  | Self employed | 1.10 (0.99, 1.20) | **1.23** (1.05, 1.41) | 1.20 (0.96, 1.48) |
|  | Student (15+) | **1.16** (1.05, 1.27) | **1.30** (1.12, 1.47) | **1.53** (1.25, 1.84) |
|  | Unemployed, seeking | **0.82** (0.71, 0.92) | 0.82 (0.68, 1.01) | 0.93 (0.69, 1.11) |
|  | Unemployed, not seeking | 0.90 (0.78, 1.04) | 0.96 (0.76, 1.10) | 0.97 (0.69, 1.17) |
| Day of week | Weekday | 0.97 (0.95, 1.01) | 0.99 (0.95, 1.02) | 0.99 (0.94, 1.04) |
|  | Weekend | 1.03 (0.99, 1.06) | 1.01 (0.98, 1.05) | 1.01 (0.96, 1.06) |
| Symptomatic | Yes | **1.20** (1.17, 1.23) | **1.21** (1.17, 1.26) | **1.18** (1.13, 1.24) |
|  | No | **0.83** (0.80, 0.85) | **0.82** (0.79, 0.85) | **0.84** (0.80, 0.89) |
| Using mask | Yes | **1.42** (1.38, 1.47) | **1.42** (1.37, 1.49) | **1.40** (1.32, 1.50) |
|  | No | **0.69** (0.66, 0.71) | **0.69** (0.66, 0.73) | **0.71** (0.66, 0.76) |
| Perceived infection likelihood | High | 0.96 (0.92, 1.00) | 1.00 (0.96, 1.05) | 1.05 (0.99, 1.12) |
|  | Neutral | 1.00 (0.97, 1.03) | 1.00 (0.96, 1.03) | 1.00 (0.93, 1.05) |
|  | Low | 1.04 (1.00, 1.08) | 1.00 (0.96, 1.04) | 0.96 (0.90, 1.02) |
| Perceived personal COVID-19 severity | High | **0.93** (0.89, 0.96) | **0.89** (0.85, 0.94) | **0.87** (0.82, 0.94) |
|  | Neutral | 0.97 (0.93, 1.01) | 0.98 (0.93, 1.02) | 0.98 (0.92, 1.04) |
|  | Low | **1.11** (1.07, 1.15) | **1.14** (1.09, 1.20) | **1.17** (1.09, 1.25) |
| Gov. advice | Trust | **1.12** (1.08, 1.16) | 1.05 (1.00, 1.10) | 1.02 (0.97, 1.09) |
|  | Neutral | **0.89** (0.86, 0.93) | **0.93** (0.87, 0.98) | **0.91** (0.84, 0.98) |
|  | Distrust | 1.00 (0.95, 1.03) | 1.02 (0.97, 1.09) | 1.07 (0.99, 1.16) |
| Age group* | 0–4 | **1.28** (1.16, 1.41) | **1.51** (1.31, 1.72) | **1.92** (1.58, 2.29) |
|  | 5–9 | **1.40** (1.27, 1.53) | **1.79** (1.56, 2.03) | **2.65** (2.22, 3.10) |
|  | 10–14 | **1.36** (1.24, 1.48) | **1.68** (1.48, 1.89) | **2.58** (2.18, 3.02) |
|  | 15–19 | 0.96 (0.86, 1.07) | 1.13 (0.98, 1.28) | **1.24** (1.04, 1.47) |
|  | 20–24 | **0.74** (0.67, 0.82) | **0.72** (0.62, 0.82) | **0.68** (0.56, 0.81) |
|  | 25–34 | **0.74** (0.69, 0.80) | **0.65** (0.58, 0.72) | **0.57** (0.49, 0.65) |
|  | 35–44 | **0.88** (0.82, 0.94) | **0.81** (0.73, 0.89) | **0.62** (0.53, 0.71) |
|  | 45–54 | **0.86** (0.80, 0.91) | **0.85** (0.77, 0.92) | **0.69** (0.60, 0.78) |
|  | 55–64 | 0.95 (0.90, 1.01) | **0.91** (0.83, 0.99) | **0.78** (0.69, 0.89) |
|  | 65–69 | 0.98 (0.92, 1.04) | 0.94 (0.85, 1.03) | **0.82** (0.72, 0.94) |
|  | 70–74 | 1.06 (0.98, 1.14) | **0.82** (0.72, 0.93) | **0.68** (0.55, 0.85) |
|  | 75–79 | 0.93 (0.82, 1.05) | **0.79** (0.62, 0.95) | 0.77 (0.53, 1.04) |
|  | 80–85 | 1.07 (0.88, 1.26) | 1.05 (0.75, 1.36) | 1.04 (0.62, 1.57) |
| Sex* | Male | **0.96** (0.94, 0.98) | 0.99 (0.96, 1.02) | 1.02 (0.98, 1.07) |
|  | Female | **1.04** (1.02, 1.07) | 1.01 (0.98, 1.05) | 0.98 (0.93, 1.02) |
| Participation* | First-time | **1.17** (1.13, 1.20) | **1.23** (1.18, 1.28) | **1.27** (1.21, 1.34) |
|  | Repeated | **0.85** (0.83, 0.88) | **0.81** (0.78, 0.84) | **0.78** (0.74, 0.83) |

\*Age group, Sex, and Participation are treated as control variables; no variable selection is performed.
<sup>1</sup>RR 20%: Relative risk (posterior median with 95% CrI) of having daily non-household contacts in the top 20% of the population within the given exposure group, compared to the population average. Bold values indicate statistical significance at the 95% credible level (i.e., the 95% CrI does not include 1). Red (blue) values indicate statistically significant positive (negative) associations.

Individuals living alone were substantially more likely to be in the high-contact tail than those in larger households. For example, living in a one-person household was associated with a 20% higher risk of being in the top 20% contact group compared to the population average, whereas living in a household of five or more reduced that risk by about 14%. This suggests that people who live alone compensate by seeking more contacts outside the home, while those in large households satisfy more of their social needs within the household, especially under pandemic-related pressure to reduce external contacts. Employment status was another strong predictor. Full-time employees and students had markedly elevated risks of appearing in the top 10% and 5% of the contact distribution, reflecting the contact intensity of workplaces and educational settings. In contrast, full-time parents and retirees were strongly under-represented in the tail. Attitudinal and behavioural variables also mattered. Individuals who perceived COVID-19 as a serious personal threat were less likely to be in the highest contact groups, while those with low concern were more likely to appear there. Mask use showed an apparent paradox: mask wearers were more likely to be in the tail than non-wearers. This likely reflects risk compensation though the broader evidence, including from video observational studies, remains mixed. ^32^ Similarly, and consistent with higher infection risk among people with high exposure metrics, ^33,34^ reporting recent COVID-like symptoms was also associated with tail membership. For pandemic time periods, these findings highlight that a wider set of tailored interventions beyond just reducing mass gatherings will need to be in place to control super-spreading and allow easing population-wide restrictions.

### Heavier tails are associated with higher incidence

Substantial empirical data supports the social contact hypothesis that higher social contact intensities precede and are linked to higher disease incidence, ^35–37^ however data supporting the role of heavier tails on population-level transmission is less clear. Germany’s second wave was spatially heterogeneous, with three distinct regional patterns of incidence emerging during late 2020 and early 2021 (Fig. 3a-b). ^28^ To test the contact tail hypothesis, we overlaid the COVIMOD data on this classification by assigning each survey respondent to their corresponding group, and analysing contact patterns for November 2020–April 2021. The contact distributions differed systematically across the three region groups not only in mean numbers, but crucially the higher incidence Group 2 also exhibited significantly heavier tails in their degree distribution (Fig. 3c). These results suggest that regional differences in incidence during the second wave were not solely due to differences in average contact rates or initial conditions. Instead, the underlying heterogeneity of contact patterns—captured by the heaviness of the tail—appears to have played a key role, in line with theoretical expectations ^7,9^ and individual-level data showing that higher exposure measures were associated with increased infection risk. ^33,34^

**Fig. 3.**
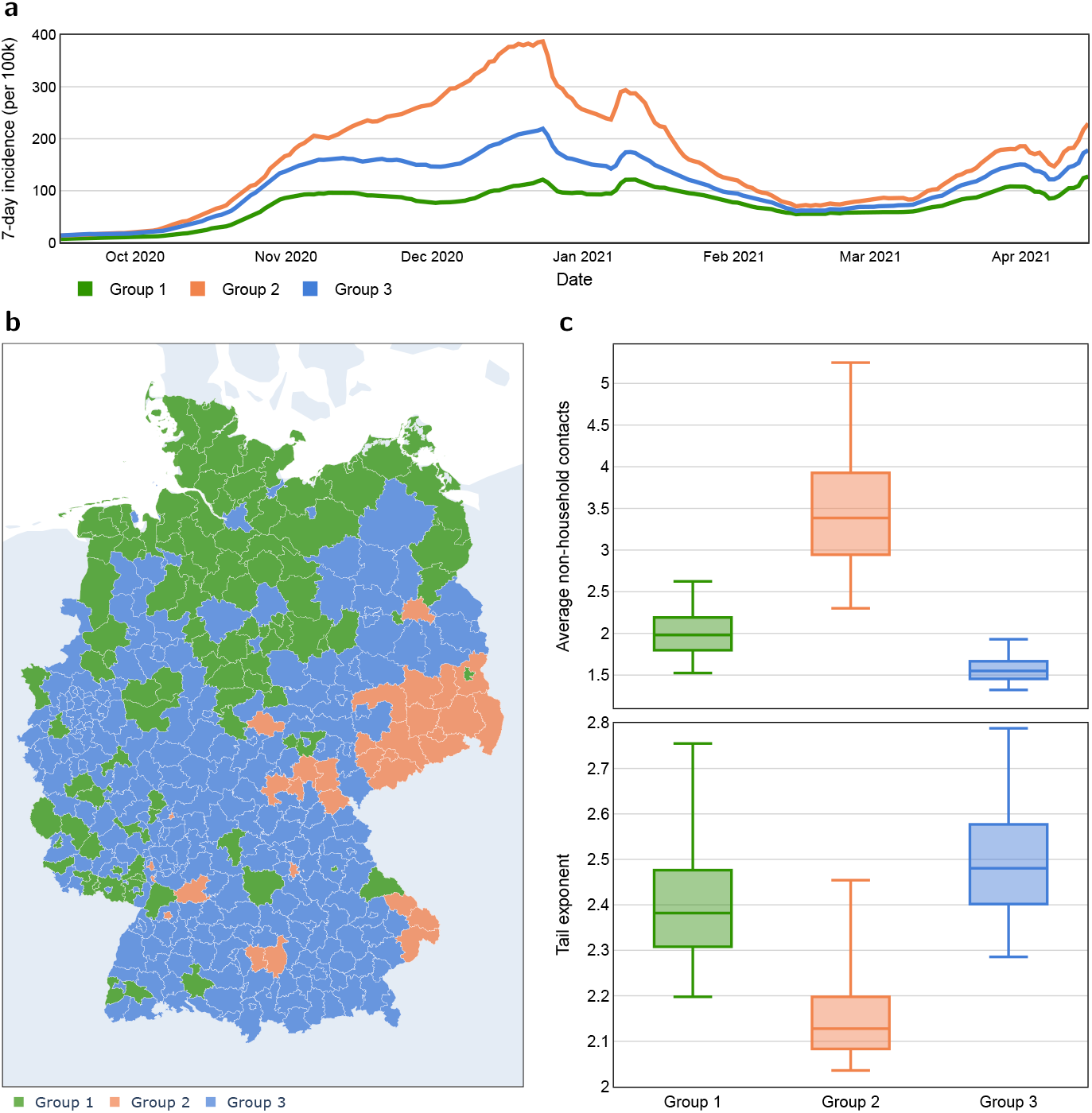
Variation in transmission dynamics parallels regional differences in contact patterns. **a**, Weekly incidence of reported SARS-CoV-2 cases (per 100,000 inhabitants) for the three region groups. b, German NUTS 3 regions are clustered into three groups based on the similarity of transmission patterns observed from September to December 2020. c, Contact patterns across the three groups during the second lockdown (November 2020 to April 2021). Upper: Boxplots of average daily non-household contacts (posterior median, IQR, and 95% CrI). Lower: boxplots of the tail exponent of the corresponding contact network (bootstrapped median, IQR, and 95% CI).

### Tailoring epidemic interventions to pandemic contact networks

In networks characterised by heavy-tailed degree distributions, transmission is disproportionately driven by high-degree nodes: infections are more likely to “burn through” connected individuals early, producing faster initial spread than random-mixing models predict. ^39^ In contrast, random networks with static structure show reduced early growth rates and final epidemic sizes compared to random mixing models ^40^ (Supplementary Fig. 3). This concentration of connectivity also creates opportunities for interventions tailored to protect high-contact individuals and thereby disrupt transmission chains (Fig. 4a). ^7,9^

**Fig. 4.**
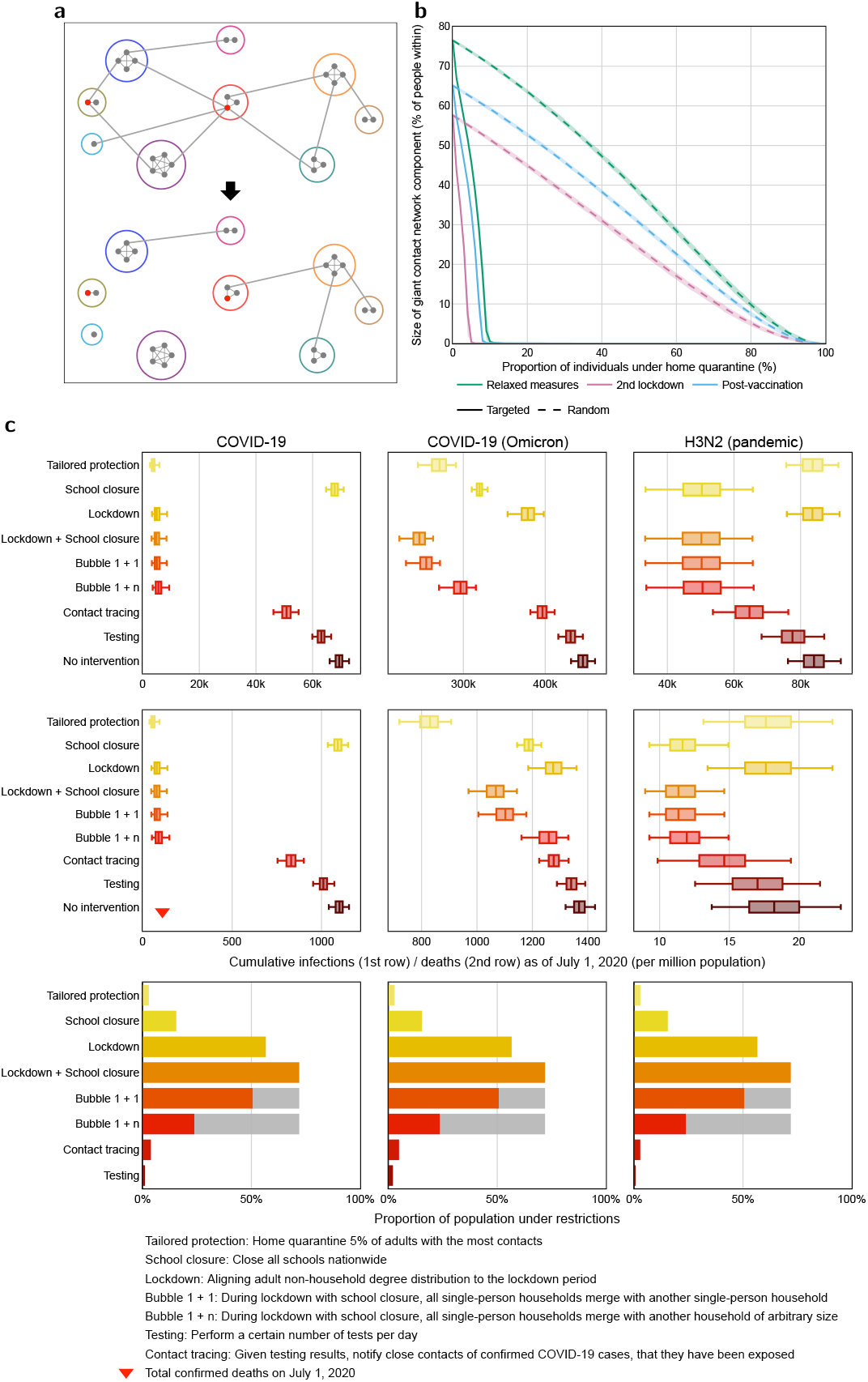
Modelling the effect of tailored intervention on network structure and epidemic spread. **a**, Diagram of a community contact network with household clusters (coloured circles) before (top) and after (bottom) protective intervention for the two individuals with the most non-household connections (red solid dots). b, Giant component proportion in a synthetic German contact network versus the proportion of the population under intervention (lines: average in 1,000 simulations, shaded bands: 95% CIs). The plot compares a structurally-tailored intervention prioritising individuals with the most non-household contacts (solid lines) against an untailored intervention (dashed lines) under three pandemic phases (distinguished by colour). d, Agent-based simulations on a synthetic multilayer contact network for three pathogens: COVID-19 (ancestral strain), COVID-19 (Omicron) and influenza A (H3N2). For each pathogen, nine intervention strategies are evaluated. Boxplots (median, IQR, 95% CI) show the distribution of cumulative infections (1st row) and cumulative deaths (2nd row) per million population by 1 July 2020. The scale of implementation for each intervention scenario (3rd row). The bars quantify the percentage of agents subject to restrictions (lockdown/isolation/quarantine) in one typical simulation. The gray part of the bar represents the proportion of individuals covered by the bubble policy.

We assessed this theoretical principle in the context of Germany’s age-specific population, household structure, and our empirically estimated non-household contact distributions for three template pathogens: the original SARS-CoV-2 strain, the more infectious but less severe Omicron variant, and a hypothetical pandemic influenza A (H3N2) strain with low disease severity but higher susceptibility and transmissibility among children. Specifically, we considered a stylised community network comprising 3.3m agents (4% of Germany’s population), in which households are parameterised according to census characteristics and form fully connected cliques. Transmission rates per contact within household are set to be higher than in other layers for each pathogen, with values roughly aligned with best available estimates to reflect the longer and closer contact between household members. Individuals maintain non-household links as per our estimated non-household degree distributions for each phase. We then sequentially “removed” an increasing fraction *p* of nodes either in descending order of degree (structurally-tailored intervention) or at random, and tracked the size of the largest connected component.

Percolation theory predicts a critical threshold *p*_*c*_ at which the giant component collapses. ^41^ We found that Germany’s synthetic contact network retained a low percolation threshold in any of the five phases when individuals were removed at random, but was highly sensitive to structurally-tailored interventions (Fig. 4b). This substantiates the potential of tailoring interventions to high-contact individuals for epidemic control, leveraging the actual shape of Germany’s non-household degree distribution. For example, in the relaxed measures phase (June–September 2020), interventions reaching ∼ 10% of the most connected individuals was sufficient to reduce the giant component to *<* 1% of the population, whereas random removal (untailored interventions) required eliminating well over 90% of individuals to achieve the same effect.

To translate these structural insights into epidemic outcomes, we simulated the spread of the three template pathogens on the synthetic multilayer networks with the agent-based Covasim framework, ^8^ incorporating realistic household, school, and combined work and community layers and pathogen-specific transmission parameters (details in Supplementary Information). All simulations began on 1 February 2020, with 0.1% of randomly selected individuals initially infected.

We compared several intervention scenarios (Fig. 4). The *testing* scenario implemented isolation of people 1 day after they tested positive, and we parameterised testing capacity via the empirical number of tests conducted in Germany. Akin to digital contact tracing and notification services, ^34^ our *contact tracing* scenario expanded testing through tracing a proportion of household, school, and combined work and community contacts (100%, 50%, and 30%, respectively), with immediate quarantine of traced contacts for 14 days. The *lockdown* scenario, initiated once 500 deaths were detected nationwide, and then contacts in the combined work-community layer were set to match Germany’s low-degree distribution observed in the first lockdown phase. Additional school closures were considered in the *lockdown+school closure* (LS) scenario. Two bubble scenarios relaxed LS: in the *Bubble 1+1* intervention, all adults living alone formed exclusive pairwise bubbles with another person living alone; in the *Bubble 1+n* policy, each single-person household formed a bubble with one other arbitrary household. The *tailored protection* scenario also initiated after detecting 500 deaths, and then the top 5% most connected adults received priority interventions including preventive home quarantine; schools were not closed.

For a pathogen as the original SARS-CoV-2 variant, the tailored protection, lockdown and LS scenarios produced large and comparable reductions in cumulative infections and deaths by mid-2020 relative to no intervention (Fig. 4c, left column). Remarkably, providing protective interventions to just 5% of adults with the highest degrees matched the impact of a nationwide lockdown with school closures, while disrupting far fewer people. For a comparatively more transmissible but less severe pathogen like Omicron, tailored protective interventions outperformed all other modelled interventions: focusing on the most connected 5% averted 40% (30%-50%) more deaths compared to no intervention in our synthetic population 5 months after. In contrast, considering a pathogen such as pandemic influenza A (H3N2) that is more easily spreading through children compared to adults, school closures were essential, and tailored protective interventions were not the optimal intervention scenario. The bubble policies sat between these extremes. Bubble 1+1 maintained substantially reduced epidemic sizes relative to lockdown with school closure, though the impact of Bubble 1+n was more modest for the more highly transmissible Omicron. Testing alone slowed but did not prevent widespread transmission, and test-and-trace never matched the beneficial impact of interventions that directly addressed the tail of the degree distribution, even assuming immediate tracing upon infection and full compliance of traced individuals. Overall, these results indicate that in real-world heavy-tailed contact networks and across all pandemic phases, interventions that specifically support and protect high-contact individuals can often control epidemics as well as population-wide interventions in terms of cumulative infections and deaths averted, and more effectively so when the number of individuals impacted by the intervention are considered; childhood-driven infectious diseases remain an important exception.

## Discussion

Our integrated analysis of nationwide, longitudinal non-household contact survey data, novel statistical heavy-tail models and agent-based epidemic intervention counterfactuals shows that throughout the COVID-19 pandemic, Germany’s non-household contact network remained heavy-tailed. Crucially, the individuals who sustained this heavy tail were not classic superspreaders attending concerts, sporting events, or large gatherings—the top 5% quantile had just 9.4 non-household contacts per day. Instead, these were predominantly working-age adults whose elevated contact rates stemmed from occupational necessities: full-time employees, students, and those in public-facing roles whose jobs inherently require in-person interaction. Investigations from across Europe suggest that these findings generalise beyond Germany. ^43–46^ Beta-Negative-Binomial distributions provided the best fit to the non-household contact data across all five pandemic phases. Across these, spanning two lockdowns and intervening relaxations, the most connected ∼ 20% of individuals consistently generated more than 80% of all reported non-household contacts, a phenomenon widely known as the Pareto principle. ^47^ Apart from the first lockdown, the best-fitting degree distribution was characterised by infinite variance implying that theoretically, pathogens with arbitrarily small transmission risk could have continued to spread across the network, ^7,9^ though in practice these theoretical predictions are limited by finite population size, modularity, and saturation effects. ^30,48^ The primary implications are: early epidemic growth ignites rapidly with high attack rates if an index case reaches a social hub early; ^7,30^ super-spreading is frequent rather than exceptional; ^1^ epidemic control can be achieved more efficiently by targeting hubs and otherwise attack rates flatten naturally when hubs become immune; ^49^ social inequality that entrenches hubs, connectivity and crowding becomes a driver of spread; ^18,50,51^ and epidemics are likely to persist and become endemic. ^7,9^

By fitting heavy-tail models to comprehensive contact survey data and evaluating agent-based counter-factual intervention scenarios, our work bridges a gap between theoretical models and empirical observations. Many network-based epidemic models assume scale-free degree distributions or strong contact heterogeneity in order to reproduce superspreading, low epidemic thresholds and complex outbreak dynamics. ^52–58^ Direct evidence for such behaviour at the population level has been scarce, ^44^ particularly under the influence of substantial, broad-ranging NPIs. Here we provide such evidence for Germany, and show that heavy tails persisted throughout the pandemic, but also that the entire contact network shifted temporarily towards a finite-variance regime during the first lockdown. This “structural plasticity” underscores that population-wide NPIs can indeed do more than scale down average contacts: they may alter the entire network topology and eliminate infinite-variance regimes associated with highly stochastic resurgence, entrenched inequity in disease spread, and epidemic persistence, though typically at high societal and economic costs. ^59^

We also identify who populated the high-contact tail during the COVID-19 pandemic. Contrary to assumptions implicit in policies that emphasise mass gathering restrictions, the tail did not correspond to a tiny set of elite hubs with 50–200 non-household contacts per day—concert-goers, nightclub patrons, or sports fans. Rather, high-contact individuals were characterised by ordinary occupational circumstances: full-time employment, student status, and living arrangements that necessitated seeking social contact outside the home. This distinction has profound policy implications. Restricting mass events, while symbolically significant and occasionally effective for specific outbreaks, ^60–62^ addresses only a fraction of the heavy tail. The bulk of high-contact individuals are workers whose contact patterns are shaped by job requirements—retail staff, healthcare workers not captured by special protocols, educators, and service-sector employees—rather than discretionary social choices.

These findings suggest that interventions tailored to the structure of contact networks should prioritise safeguarding occupational and structural contexts, including workplace ventilation, reduced crowding in public-facing settings, and flexible working arrangements, rather than focusing predominantly on mass events. ^59^ Such tailored approaches could harness venue or occupational proxies, ^63,64^ acquaintance sampling, ^49^ real-time point-of-interest hotspot mapping, ^65,66^ or venue repertoire characterisation. ^67^

Our network simulations show that recognising and acting on this structure has practical consequences. Across pandemic phases on Germany’s calibrated synthetic contact network, and not just theoretical models, ^7,30,52^ uniformly reducing contacts is structurally inefficient: substantial effort is spent suppressing already low-risk connections while high-degree individuals—predominantly essential workers—may remain exposed. By contrast, interventions tailored to protect those with occupationally-driven high contact rates can fragment transmission networks with far less societal disruption. In our simulations, structurally supporting the top 5% most connected adults through measures such as priority testing, workplace safety protocols, or flexible arrangements controlled ancestral SARS-CoV-2 at least as effectively as a lockdown with school closures, and more effectively for a highly transmissible Omicron-like variant, while disrupting far fewer lives. Bubble policies that capped each person’s number of non-household partners approximated this effect by design, turning potential hubs into members of small clusters. The comparison across pathogens highlights when tailoring interventions to contact structure matters most. For childhood-driven pathogens (such as our pandemic influenza A (H3N2) scenario with relatively higher susceptibility and transmissibility among children), even substantial contact heterogeneity did not translate into large differences between tailored and uniform NPIs, and childhood-driven epidemics were most effectively controlled through school closures. In contrast, for pathogens with higher *R*_0_ and greater transmission potential via adults—such as SARSCoV-2 and related coronaviruses ^6,68^—the network tail becomes pivotal. Because this tail comprises workers with job-driven contact patterns rather than discretionary social behaviour, interventions that support safe working conditions for high-contact occupations can reduce transmission while preserving economic activity and livelihoods. Such occupationally-tailored interventions function as structural insurance: they are most impactful precisely when the pathogen is capable of rapid spread through essential in-person workforce networks.

This study has limitations. The COVIMOD data derive from self-reported retrospective diaries, which are subject to recall error. Participants were recruited through an online panel, which can deviate substantially from the underlying population. ^69^ This could have introduced selection bias if individuals who were more compliant with public health advice were more likely to participate, and could have led to underestimation of absolute contact rates and tail heaviness. ^70^ Repeat participation is also vulnerable to survey fatigue, ^65^ though our key findings—including the persistence of heavy tails, the classification of most phases in the infinite-variance regime, and the relative performance of interventions in counterfactual simulations—were robust in sensitivity analyses conducted on first-time participants only (Supplementary Fig. 4 and Supplementary Table 2). Our contact definition, while standard in the field, ^18,27^ does not capture all interactions relevant for respiratory transmission such as short unmasked encounters in crowded public transport; however group contacts could be reported from wave 3 onwards and mitigate these concerns. Parents or guardians also reported contacts on behalf of dependent children as in previous work, ^18,19^ which has been associated with underestimation of their contacts. ^28^ We therefore excluded proxy-reported non-household contacts in sensitivity analyses, and found that our key findings remained qualitatively unchanged (Supplementary Fig. 5 and Supplementary Table 2). To alleviate this limitation, our pandemic counterfactuals also relied for children and adolescents on external data on contact rates, instead of our COVIMOD survey data. Considering that prior selection may affect Bayesian inference, we examined multiple prior settings and confirmed that the main conclusions remain consistent (Supplementary Fig. 6 and Supplementary Table 3). Finally, our counterfactuals were done on a synthetic population that matched Germany’s household structure, nonhousehold degree distributions, and school, workplace and community-level transmission settings (Methods), but lacked additional realism such as mobility, ^72^ and it remains hard to anticipate the implications of such and further realism on scenario analyses.

Despite these caveats, our results provide a coherent picture. Heavy-tailed non-household contact networks persisted in Germany throughout the pandemic, including during phases in which *R*_*t*_ was near or below one. NPIs modulated but did not consistently eliminate the tail, and the individuals who constituted the tail during the pandemic were characterised by everyday demographic and behavioural activities. These structural facts matter: they explain how epidemics can remain poised on a knife-edge even when population-wide average reproduction numbers look favourable, and reveal that interventions tailored to the contact structure of everyday working life can have outsized effects on epidemic outcomes. Designing policies that explicitly account for heavy tails in contact patterns—recognising that these tails reflect occupational necessity rather than exceptional behaviour—offers a route towards epidemic control that is both more precise and potentially more equitable than repeated population-wide lockdowns. Setting-based approaches (improved workplace ventilation, reduced crowding in service environments, hybrid working options), occupational protections (priority testing and vaccination for high-contact workers), and digital tools (venue check-ins, point-of-interest hotspot mapping) can together mitigate transmission through essential workforce networks while preserving livelihoods and social function for a broad range of infectious diseases.

## Data Availability

All data produced in the present study are available upon reasonable request to the authors.

## Methods

### Data

The COVIMOD study is a longitudinal study conducted in Germany, consisting of 33 waves from April 2020 to December 2021. Participants were recruited by the market research company Ipsos from members of the online panel i-say.com, ^73^ using quota sampling based on age, gender, and region. A subset of adult participants with children under 18 was invited to report as proxies for their children to collect data on minors. New participants were recruited when participant numbers fell below quotas due to dropout.

The COVIMOD questionnaire was based on the CoMix study. ^21,74^ A copy of the questionnaire is available in Additional File 1 of.Tomori et al. ^22^ Participants were asked to retrospectively report information on each social contact between 5 AM the previous day and 5 AM on the survey day. Contact was defined as people with whom participants met in person and with whom they exchanged at least a few words or had physical contact. Participants reported the age and gender of contacts, their relationship, contact duration, and setting (e.g., home, school, workplace). The questionnaire also covered respondents’ demographics, preventive behaviors (e.g., whether they wore a mask the previous day), and attitudes toward COVID-19 and government measures. ^24,75^ Starting from wave 3, participants could choose to report groups of people contacted in similar settings as group contacts, specifying their age range (*<*18, 18-64, 65+) and contact setting (work, school, and other), in case participants could not provide information for each individual separately. Data preprocessing and other auxiliary data used in this study are described in Supplementary Information.

### Bayesian heavy-tail regression

Traditional regression on conditional expectation cannot capture the tail heaviness. We address this limitation by extending multilevel regression and post-stratification (MRP) ^32^ to distribution regression settings.

Let *Y*_*i*_ ∈ {0, 1, 2, …} denote the number of non-household contacts for respondent *i* = 1, 2, …, *n*. We model *Y*_*i*_ using a BNB distribution: *Y*_*i*_ ∼ BNB(*µ*_*i*_, *σ*_*i*_, *γ*_*i*_), where *µ*_*i*_ = 𝔼 [*Y*_*i*_] is the mean, *σ*_*i*_ quantify tail-heaviness via the tail exponent *α*_*i*_, and *γ*_*i*_ is a dispersion parameter. We decompose the mean as 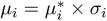, where 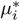 is unrelated to the tail-heaviness and *σ*_*i*_ governs tail behavior. Parameters 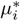, *σ*_*i*_, and *γ*_*i*_ are modelled on the log scale as

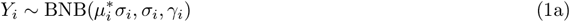

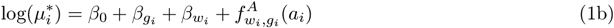

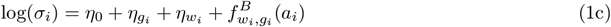

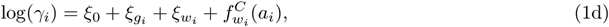

where *g*_*i*_ ∈ {*M, F* } denotes gender, *w*_*i*_ ∈ {1, …, 5} pandemic phase, and *a*_*i*_ ∈ [0, 85] age. Gender and pandemic phase were modelled with fixed effects, and age through non-parametric functions *f* for each gender and phase. All priors follow weakly informative standard recommendations.^24^

Following inference stage, we have estimated the number of non-household contacts *Y*_*g,a,w*_ for each gender *g*, age *a*, and pandemic phase *w*. We applied post-stratification weighting to obtain population-representative estimates for the distribution function *f*_pop,*w*_(*y*) and average number of non-household contacts 𝔼 [*Y*_pop,*w*_]

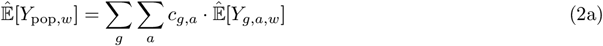

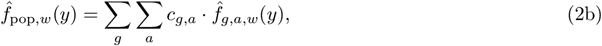

where the weights *c*_*g,a*_ were derived from the population size estimates by age and gender for 2021^5^ (table code: 12411-0041). For the tail exponent, we employ a bootstrap aggregating method: we draw *B* datasets from the posterior predictive distribution, each of size *n* and matched to the German age-gender distribution. Collecting posterior samples from each dataset yields estimates of *α*_pop,*w*_. To estimate geographic differences in contact distributions, we extend the model to include regional covariates based on the classification proposed by Schuppert et al. ^28^ .

### Bayesian variable selection

We conducted a feature selection analysis to identify characteristics associated with high-degree nodes. We assign each respondent a binary variable indicating whether their number of non-household contacts was in the tail, 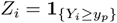, where *y*_*p*_ is the upper-*p*-percentile of estimated German contact distribution during the period where *i* was located (as shown in Fig. 2c), with *p* ∈ {1%, 5%, 10%}. Feature selection was performed using a standard Bernoulli model,

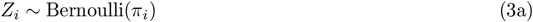

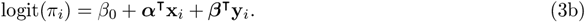

The vector of test features y_*i*_ contains one-hot encoding of the following covariates of interest: household size (1, 2, 3, 4, 5+), employment (9 categories, see Table 1), day of the week (weekday, weekend), presence of COVID-like symptoms (yes, no), whether a face mask was worn on the day of the survey (yes, no), risk perception (“I am likely to catch coronavirus”; agree, neutral, disagree), perceived severity (“Coronavirus would be a serious illness for me”; agree, neutral, disagree), and attitudes toward government advice (“If I don’t follow the government’s advice, I might spread coronavirus to someone who is vulnerable”; agree, neutral, disagree). We use sparse-inducing priors for variable selection on test features. ^33^ The vector of control features x_*i*_ includes one-hot encoding of age (13 categories, see Table 1), gender (male, female), and first-time participant status (yes, no) to control for potential confounding and reporting-fatigue effects.

### Synthetic network

We constructed a synthetic contact network of 3.3 million agents, representing the German population at a 1:25 scale. The network was structured into three standard layers: households (H), schools (S), and a combinded workplace and community layer (WC). Agents were organized into 1.7 million households, with household size distribution matching estimates for Germany in 2020^5^ (table code: 12421-0100). The S layer captured non-household contacts for individuals under 22 years of age, with a Poisson degree distribution with a mean of 11.41, based on an analysis of contact survey data from nine countries. ^34^ For the WC layer, degree sequences were sampled from our empirically estimated contact distribution.

### Transmission model

Epidemic spread was simulated using Covasim, ^8^ an agent-based modelling frame-work for COVID-19. The transmission process is described by a discrete-time compartmental model. We consider five health states: susceptible, exposed, infectious, recovered, and dead, with infected individuals further classified by symptom status (asymptomatic, pre-symptomatic, mild, severe, and critical). Agents were assigned ages corresponding to the German census (in 10-year bands), with age-dependent profiles for disease progression (e.g., susceptibility and mortality). We modelled three template pathogens: (1) ancestral SARS-CoV-2 (basic reproduction number *R*_0_ = 2.5, moderate severity); (2) Omicron variant (*R*_0_ = 9.5, lower severity); and (3) pandemic influenza A (H3N2) (*R*_0_ = 1.5, low severity, with enhanced transmissibility in children). Disease-specific parameters were drawn from published literature and provided in the Supplementary Information.

## Ethics approval

COVIMOD was approved by the ethics committee of the Medical Board Westfalen-Lippe and the University of Münster (reference number 2020-473-fs). All COVIMOD participants provided informed consent. Since this study utilized only anonymized COVIMOD data, no additional institutional review was required for this reanalysis. All analysis were carried out in accordance with relevant guidelines and regulations.

## Data availability

The subset of the COVIMOD data used in this research will be provided promptly via a secure server upon receipt of a valid request at the University of Münster. For further information, please contact Veronika Jaeger.

## Code availability

The code required to reproduce the results can be found at https://github.com/MLGlobalHealth/covimod-scale-free.

## Acknowledgements

COVIMOD was funded by intramural funds of the Institute of Epidemiology and Social Medicine, University of Münster, and of the Institute of Medical Epidemiology, Biometry and Informatics, Martin Luther University Halle-Wittenberg, as well as by funds provided by the Robert Koch Institute, Berlin, the Helmholtz-Gemeinschaft Deutscher Forschungszentren e.V. via the HZEpiAdHoc “The Helmholtz Epidemiologic Response against the COVID-19 Pandemic” project, the Saxonian COVID-19 Research Consortium SaxoCOV (co-financed with tax funds on the basis of the budget passed by the Saxon state parliament), the Federal Ministry of Education and Research (BMBF) as part of the Network University Medicine (NUM) via the egePan Unimed project (funding code: 01KX2021) and the Deutsche Forschungsgemeinschaft (DFG, German Research Foundation, project number 458526380). We acknowledge the Research Computing Service at Imperial College London (DOI: 10.14469/hpc/2232) and the National Supercomputing Centre (NSCC) Singapore (https://www.nscc.sg) for providing the high-performance computing resources used in part of this work. We thank the CoMix team and in particular Christopher Jarvis, Kevin Van Zandvoort, Amy Gimma, John Edmunds for sharing the CoMix questionnaire for the COVIMOD surveys, and for their cooperation; the team at IPSOS-Mori for their work and efforts on implementing the COVIMOD survey; Z.L. is funded by the National Medical Research Council, Singapore Ministry of Health (PREPARE-S2-2024-002); S.D. and Y.C. acknowledges funding from EPSRC Centre for Doctoral Training in Modern Statistics and Statistical Machine Learning at Imperial and Oxford (EP/S023151/1 to Prof. Axel Gandy); Y.C. acknowledges the Imperial College London President’s PhD Scholarship. S.M. acknowledges support from the National Research Foundation, Singapore, under its NRF FELLOWSHIP (NRF-NRFF15-2023-0010). The funders had no role in study design, data collection and analysis, decision to publish or preparation of the manuscript.

## Author contributions

OR conceived the initial idea for the study; ZL, OR, and SM jointly developed the study design and methodology; AK and VJ oversaw data collection; ZL, SM and OR performed the data analysis, model development, and visualization; all authors interpreted the results; SM provided the funding support; ZL, SM, OR wrote the manuscript; all authors reviewed the manuscript.

## Competing interests

The authors declare no competing interests.

## Appendix A Supplementary Infomation

### A.1 Supplementary Figures and Tables

**Supplementary Figure 1.**
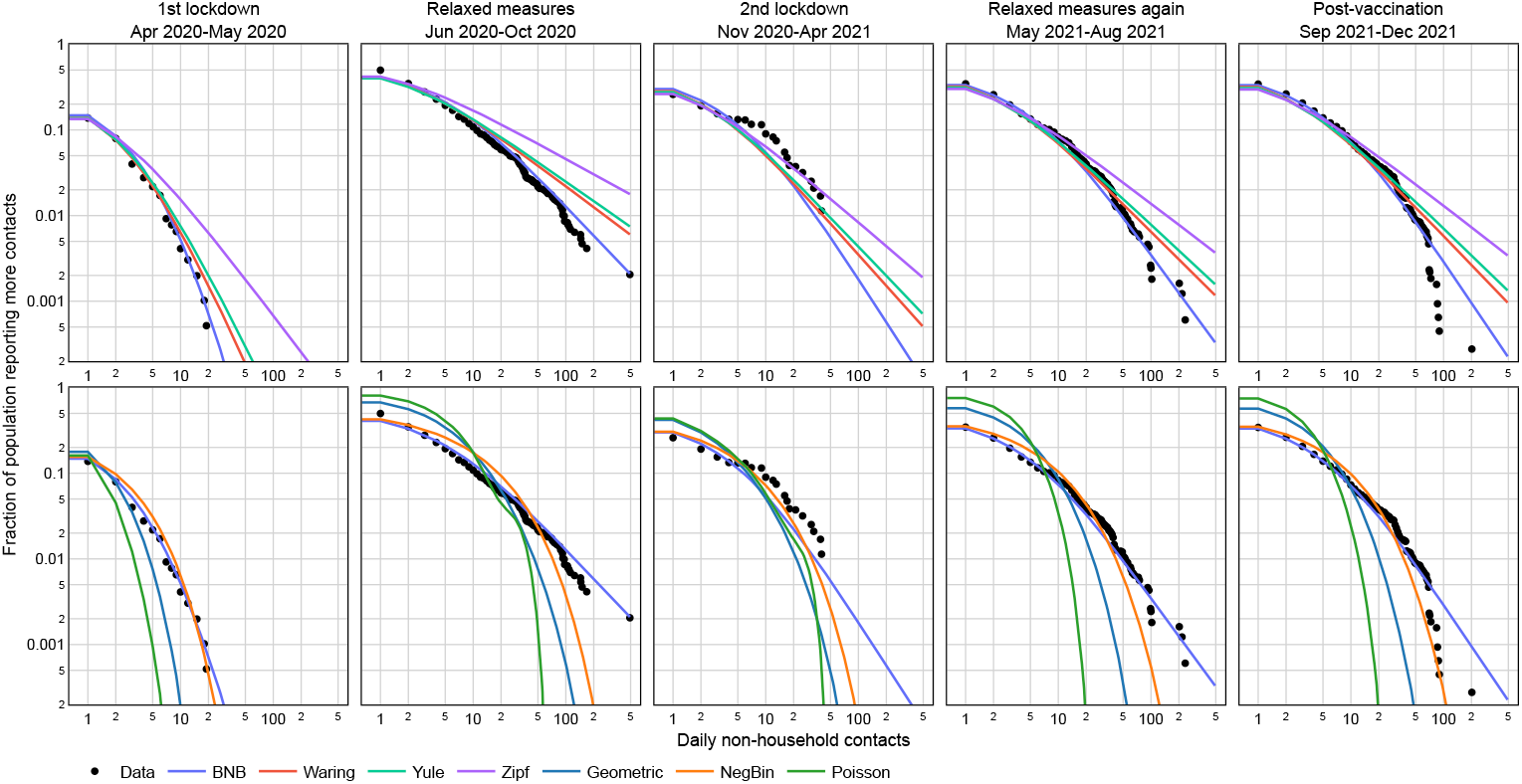
Empirical distribution of non-household contact data versus model fits across all pandemic phases. Empirical complementary cumulative distribution function (CCDF) (black dots) and posterior median CCDF of each model on log-log scale (coloured lines). Models were fitted on data from first-time participants. The top row shows heavy-tailed distribution models, and the bottom row thin-tailed distributions.

**Supplementary Figure 2.**
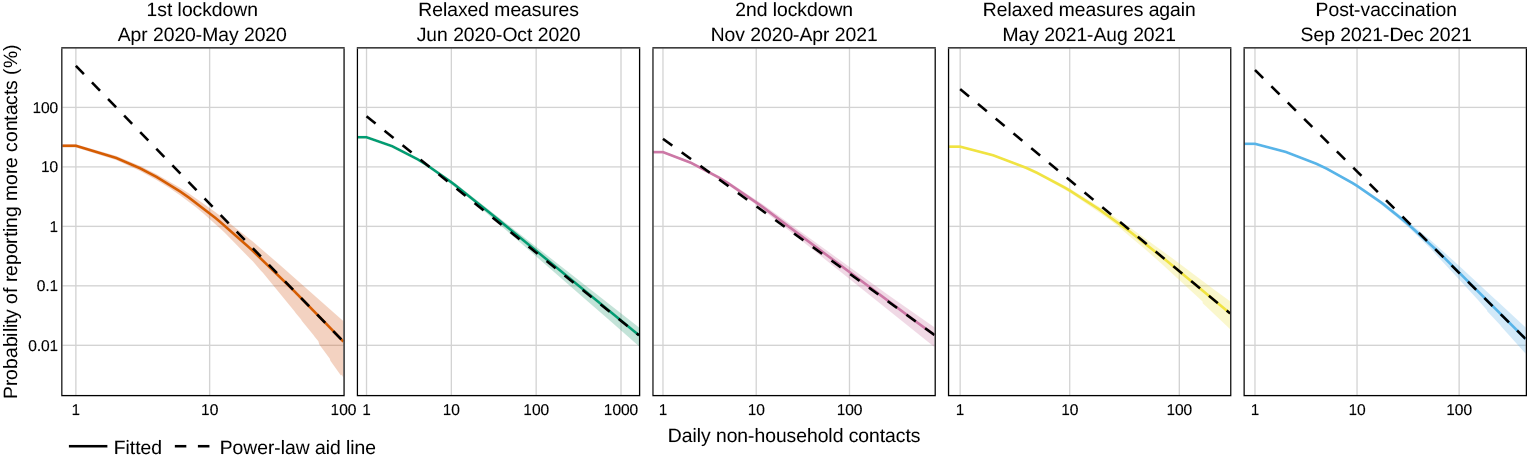
Deviations of non-household degree distributions from a pure power law across all pandemic phases. The posterior median (95% CrI) (colored solid lines and shaded areas) of the CCDF of the German non-household contact distribution across all pandemic phases. The dashed line represents a pure power law that coincides with the tail of the fitted values.

**Supplementary Figure 3.**
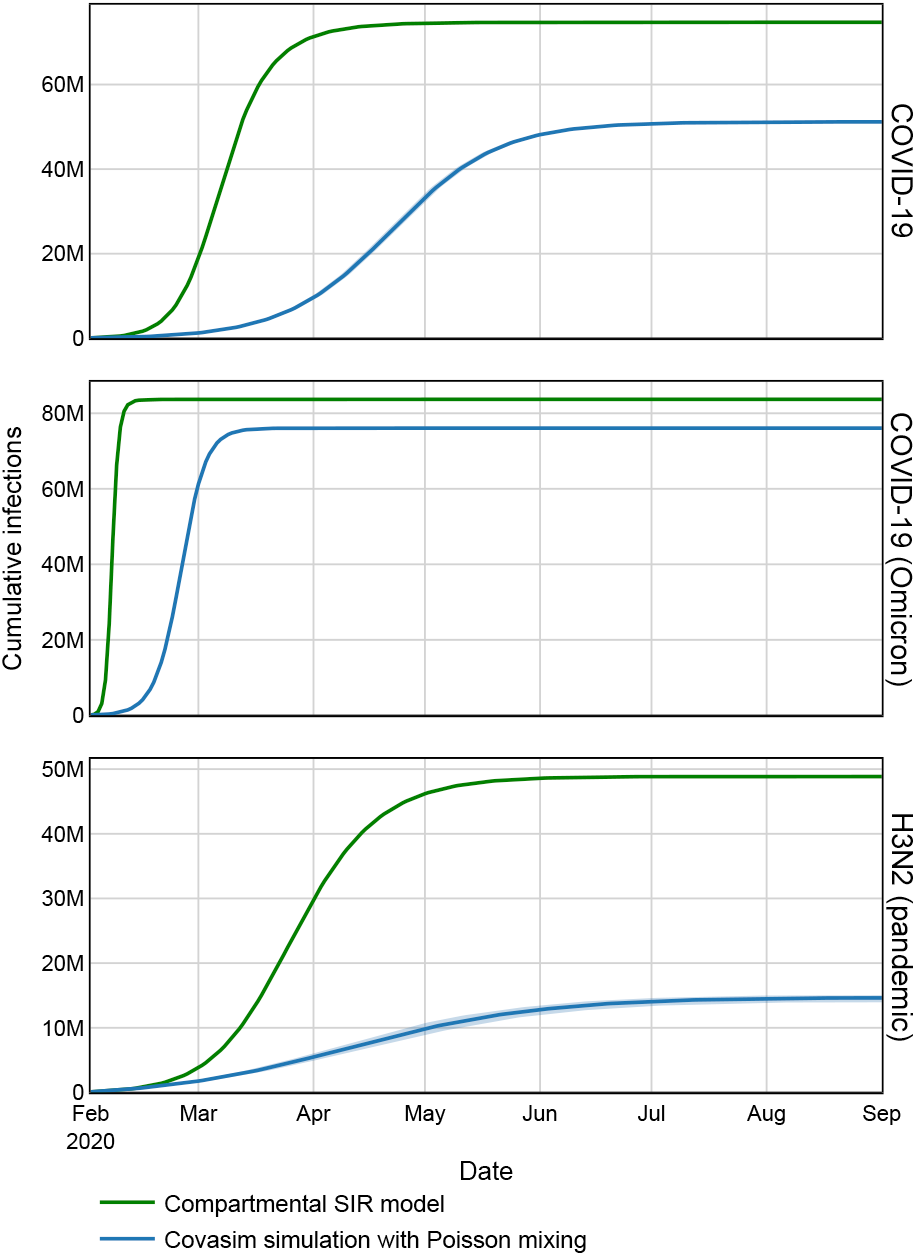
Agent-based epidemic simulations on random contact networks versus compartmental SIR (random mixing) model. We conducted agent-based epidemic simulations using the Covasim simulation software on a synthetic contact network with Poisson degree distribution of mean degree 5, and recorded cumulative infections for three template pathogens (blue). Epidemic outcomes were compared with those of a compartmenal SIR model using the same basic reproduction number *R*_0_ and mean infectious period 1*/*γ as in the Covasim simulations (green). Shown are median values and 95% empirical uncertainty intervals from 1,000 simulation runs.

**Supplementary Figure 4.**
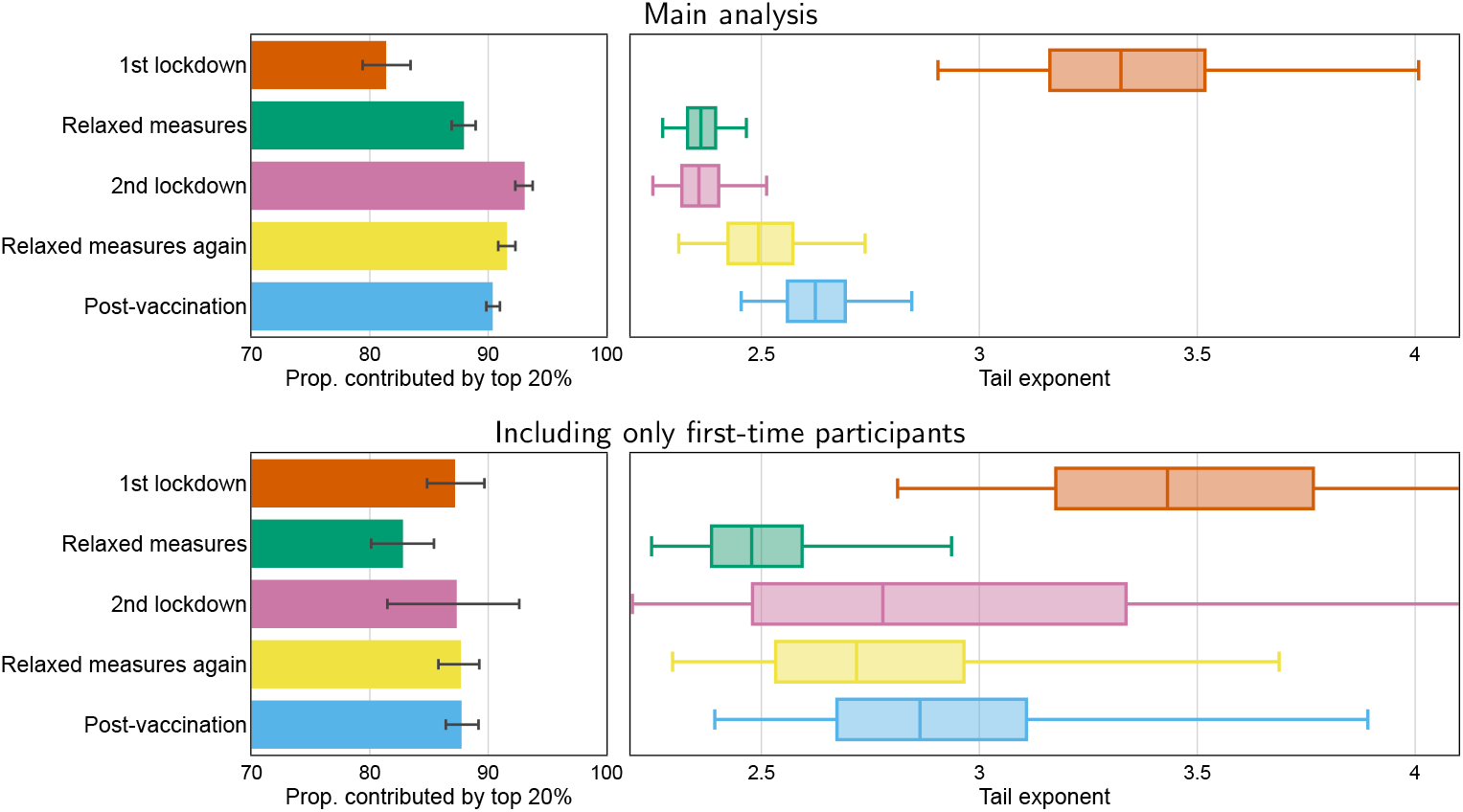
Sensitivity in heavy tail properties to repeat participation and reporting fatigue. Left: the posterior medians (bars) and 95% CrIs (error bars) for the proportion of non-household contacts contributed by the top 20% of the population. Right: tail exponent of the German contact network (bootstrapped median, IQR, and 95% CI).

**Supplementary Figure 5.**
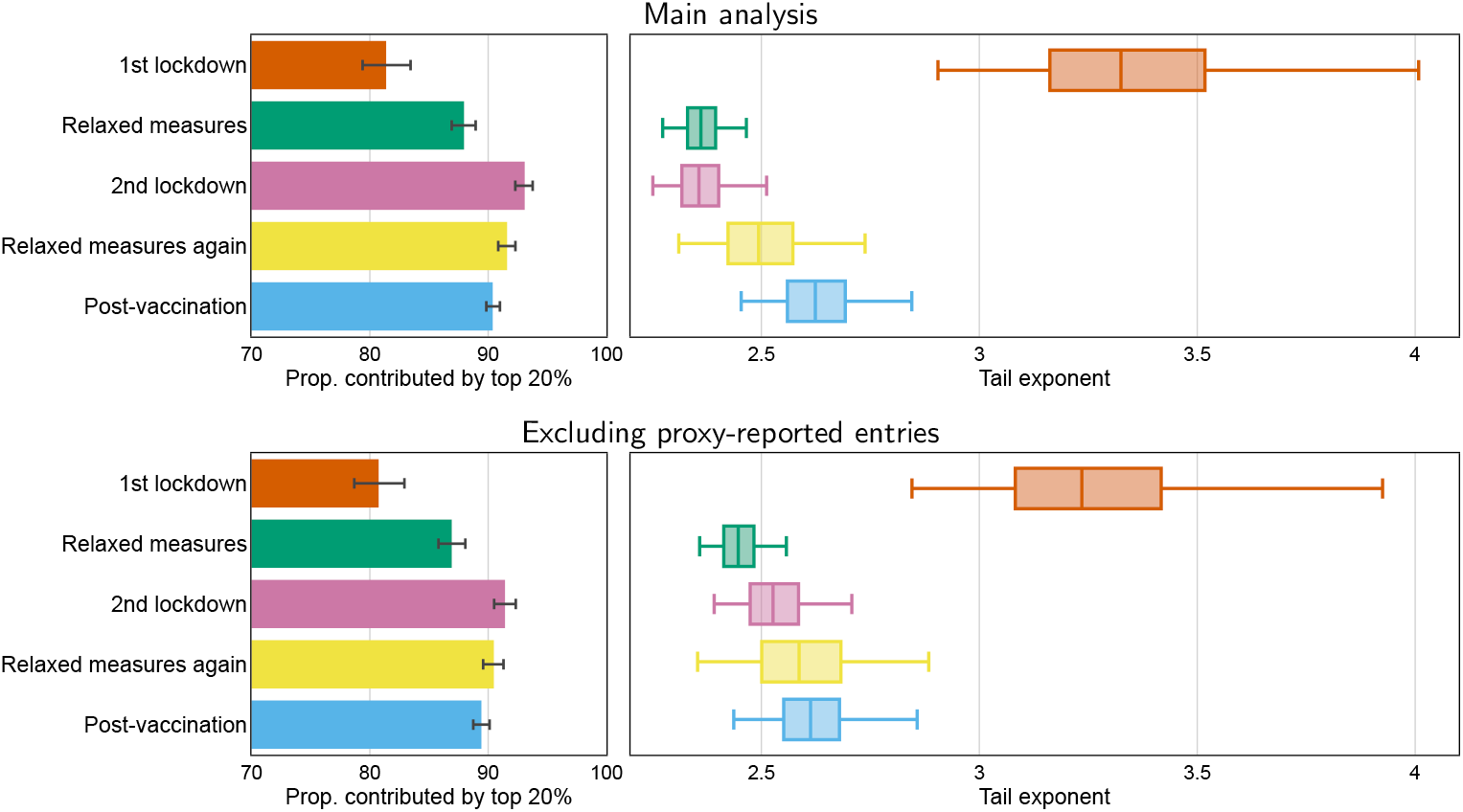
Sensitivity in heavy tail properties to proxy reporting from guardians or parents of non-household contacts of their dependent children. Left: the posterior medians (bars) and 95% CrIs (error bars) for the proportion of non-household contacts contributed by the top 20% of the population. Right: tail exponent of the German contact network (bootstrapped median, IQR, and 95% CI).

**Supplementary Figure 6.**
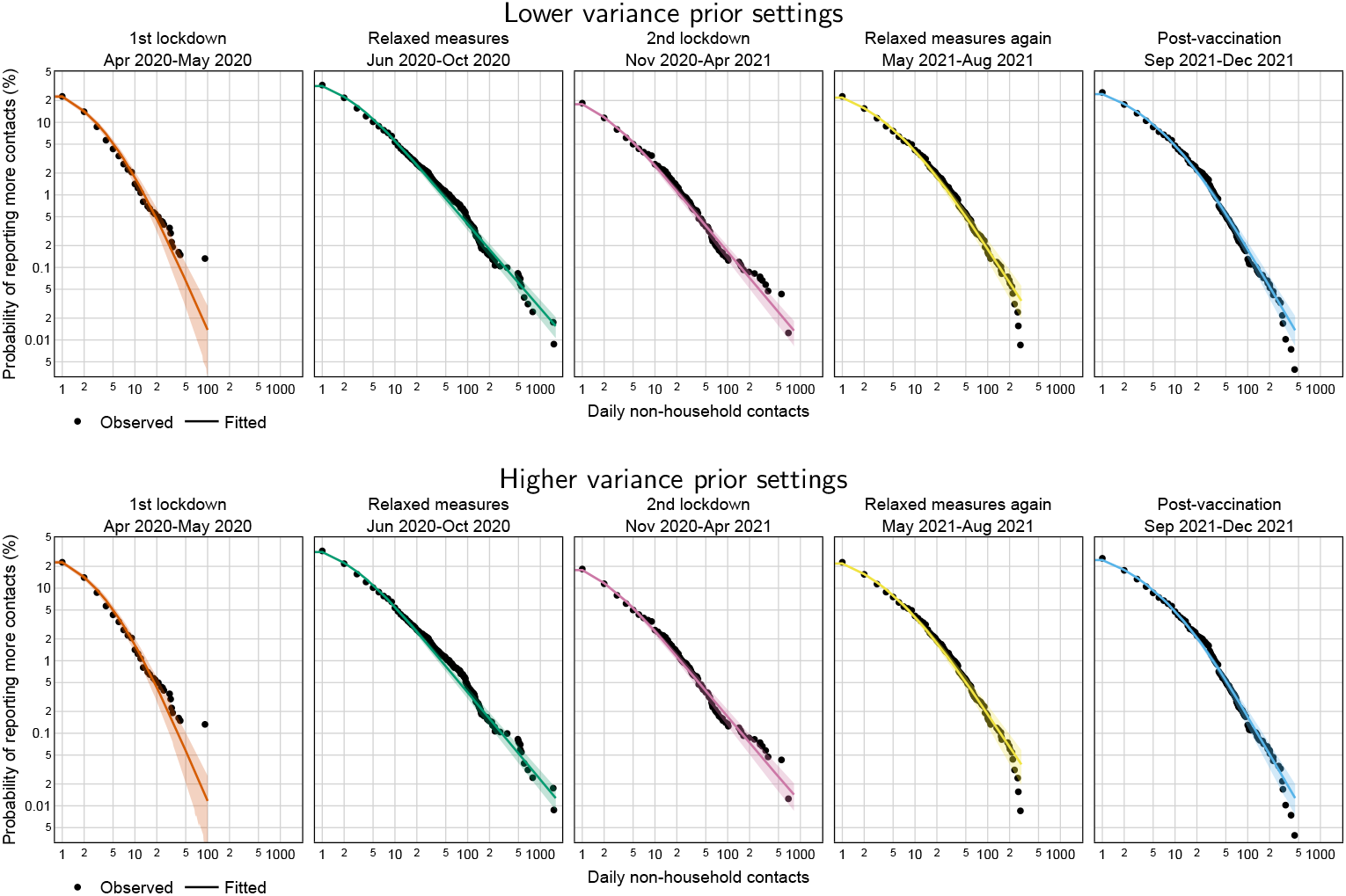
Sensitivity in the shape of population-level non-household contact distributions across pandemic phases under alternative prior specifications. Empirical CCDF of non-household contact data (black dots) versus model fits (solid lines: posterior medians, shaded areas: 95% CrIs) across all pandemic phases. Upper: lower variance prior specification; Lower: higher variance prior specification.

**Supplementary Table 1.** Model comparison of a range of count distributions for non-household contact data across all pandemic phases.

| Model | ELPD | Diff. in ELPD | SE of Diff. | Eff. P | $\hat{k}$ diagnosis |
| --- | --- | --- | --- | --- | --- |
| BNB | -14397.4 | 0.0 | 0.0 | 30.9 | Pass |
| Waring | -14443.6 | 46.2 | 8.8 | 23.7 | Pass |
| Yule | -14447.9 | 50.4 | 9.1 | 19.8 | Pass |
| Zipf | -14480.0 | 82.6 | 13.8 | 16.7 | Pass |
| NegBin | -14590.4 | 193.0 | 34.2 | 90.8 | Fail |
| Geometric | -17103.3 | 2705.9 | 152.3 | 599.1 | Fail |
| Poisson | -47716.0 | 33318.6 | 2332.7 | 3626.3 | Fail |

Model performance ranked by ELPD. The five listed metrics are: ELPD-based LOO-CV, difference in ELPD between the model and the best model (BNB), standard error of the ELPD difference, estimated effective number of parameters, and Pareto $\hat{k}$ diagnostic status. To assess whether one model outperforms another in terms of ELPD, interpret the ELPD difference with its uncertainty (SE of Diff.). Diff. in ELPD > 0 indicates worse performance than the best model. If $\Delta\text{ELPD}/\text{SE}(\Delta\text{ELPD}) > 2$ , the difference is considered statistically significant.

**Supplementary Table 2.** Sensitivity of proxy-reporting and repeat participation to the strength of association of demographic and behavioural features with the top 20% heavy tail.

| Variable | Category | Association<sup>2</sup> | Main analysis<sup>3</sup> | Exclude proxy-reported | 1st-time participants |
| --- | --- | --- | --- | --- | --- |
| Household size | 1 person | + | **1.00** | 1.00 | 1.00 |
|  | 2 person | + | **1.00** | 0.86 | 0.71 |
|  | 3 person | − | **1.00** | 1.00 | 0.88 |
|  | 4 person | − | 0.84 | 0.50 | 0.83 |
|  | 5+ person | − | **1.00** | 0.97 | 0.94 |
| Employment | Employed, full-time | + | **1.00** | 1.00 | 1.00 |
|  | Employed, part-time | + | **1.00** | 1.00 | 0.96 |
|  | Full-time parent | − | **1.00** | 1.00 | 1.00 |
|  | Long-term sick/disabled | − | 0.63 | 0.64 | 0.22 |
|  | Retired | − | 0.88 | 0.71 | 0.90 |
|  | Self employed | + | 0.98 | 0.98 | 0.84 |
|  | Student (15+) | + | **1.00** | 1.00 | 0.97 |
|  | Unemployed, seeking | − | **1.00** | 1.00 | 0.82 |
|  | Unemployed, not seeking | − | 0.92 | 0.95 | 0.70 |
| Day of week | Weekday | − | 0.98 | 0.99 | 0.00 |
|  | Weekend | + | 0.98 | 0.99 | 1.00 |
| Symptomatic | Yes | + | **1.00** | 1.00 | 1.00 |
|  | No | − | **1.00** | 1.00 | 1.00 |
| Using mask | Yes | + | **1.00** | 1.00 | 1.00 |
|  | No | − | **1.00** | 1.00 | 1.00 |
| Perceived infection likelihood | High | − | 0.99 | 0.94 | 0.76 |
|  | Neutral | + | 0.52 | 0.42 | 0.34 |
|  | Low | + | 1.00 | 0.98 | 0.88 |
| Perceived personal COVID-19 severity | High | − | **1.00** | 1.00 | 0.99 |
|  | Neutral | − | 0.97 | 0.85 | 0.29 |
|  | Low | + | **1.00** | 1.00 | 0.96 |
| Gov. advice | Trust | + | **1.00** | 1.00 | 0.99 |
|  | Neutral | − | **1.00** | 1.00 | 0.92 |
|  | Distrust | − | 0.61 | 0.28 | 0.76 |
| Age group<sup>1</sup> | 0–4 | + | **1.00** | 1.00 | 1.00 |
|  | 5–9 | + | **1.00** | 1.00 | 0.98 |
|  | 10–14 | + | **1.00** | 1.00 | 1.00 |
|  | 15–19 | − | 0.76 | 1.00 | 0.14 |
|  | 20–24 | − | **1.00** | 1.00 | 0.88 |
|  | 25–34 | − | **1.00** | 1.00 | 0.99 |
|  | 35–44 | − | **1.00** | 0.72 | 0.77 |
|  | 45–54 | − | **1.00** | 0.92 | 0.98 |
|  | 55–64 | − | 0.96 | 0.01 | 0.95 |
|  | 65–69 | − | 0.75 | 0.00 | 1.00 |
|  | 70–74 | + | 0.92 | 1.00 | 0.28 |
|  | 75–79 | − | 0.87 | 0.31 | 0.87 |
|  | 80–85 | + | 0.77 | 0.93 | 0.69 |
| Sex<sup>1</sup> | Male | − | **1.00** | 1.00 | 1.00 |
|  | Female | + | **1.00** | 1.00 | 1.00 |
| Participation<sup>1</sup> | First-time | + | **1.00** | 1.00 | 1.00 |
|  | Repeated | − | **1.00** | 1.00 | 1.00 |

<sup>1</sup> Age group, Sex, and Participation are treated as control variables; no variable selection is performed.
<sup>2</sup> The estimated direction of association (+ for positive or − for negative) in main analysis.
<sup>3</sup> Posterior probability of feature significance $\hat{p} = \max\{\hat{p}_+, \hat{p}_-\}$ , where $p_+ = \mathbb{P}(\text{RR}_j > 1 \mid D)$ , and $p_- = \mathbb{P}(\text{RR}_j < 1 \mid D)$ . Values in bold in the Main analysis column correspond to those variables found to be significant in the main analysis.

**Supplementary Table 3.**
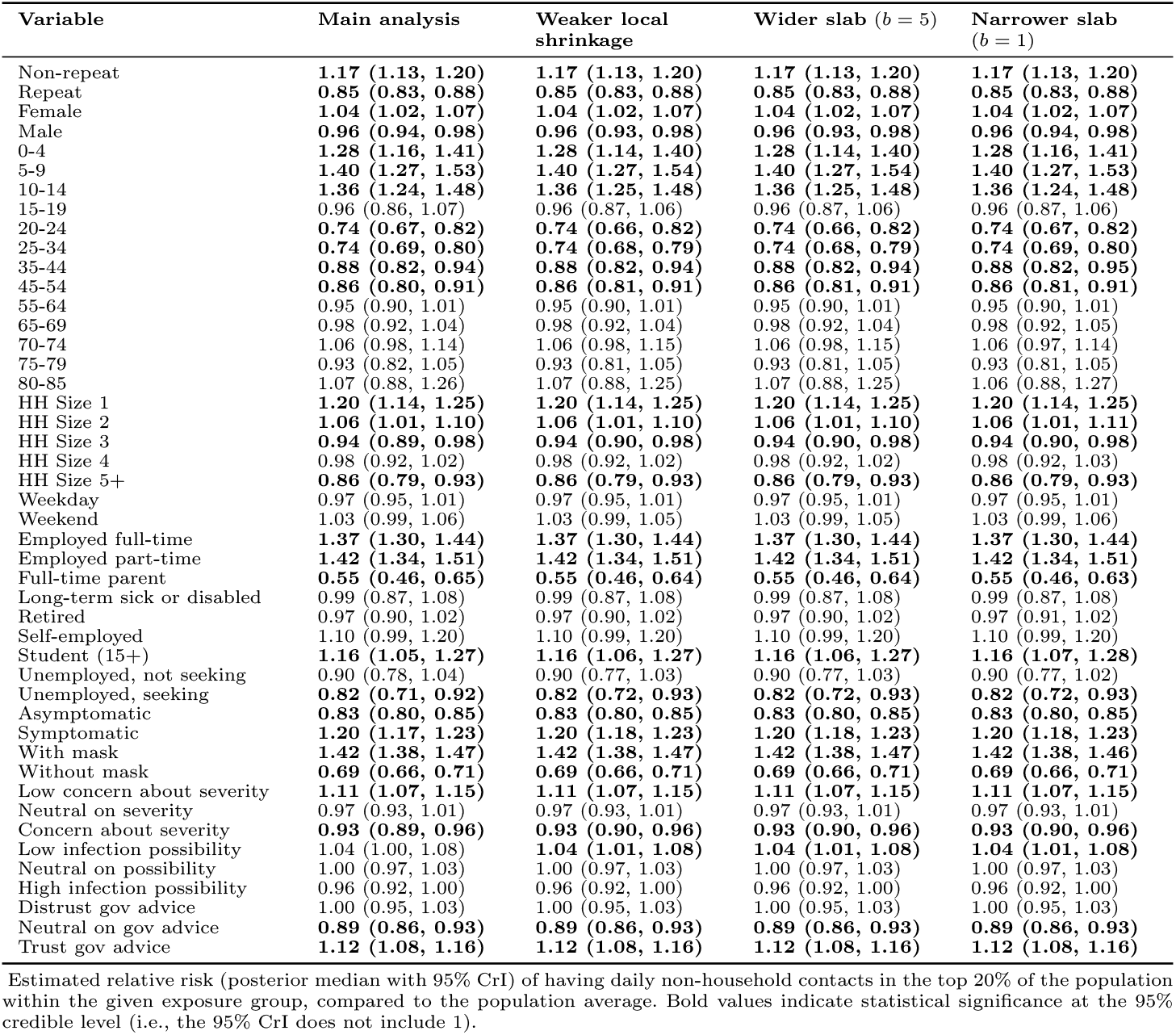
Sensitivity in associations between demographic and behavioural features with the top 20% heavy tail under alternative feature selection prior specifications.

| Variable | Main analysis | Weaker local shrinkage | Wider slab ($b = 5$) | Narrower slab ($b = 1$) |
| --- | --- | --- | --- | --- |
| Non-repeat | **1.17 (1.13, 1.20)** | **1.17 (1.13, 1.20)** | **1.17 (1.13, 1.20)** | **1.17 (1.13, 1.20)** |
| Repeat | **0.85 (0.83, 0.88)** | **0.85 (0.83, 0.88)** | **0.85 (0.83, 0.88)** | **0.85 (0.83, 0.88)** |
| Female | **1.04 (1.02, 1.07)** | **1.04 (1.02, 1.07)** | **1.04 (1.02, 1.07)** | **1.04 (1.02, 1.07)** |
| Male | **0.96 (0.94, 0.98)** | **0.96 (0.93, 0.98)** | **0.96 (0.93, 0.98)** | **0.96 (0.94, 0.98)** |
| 0-4 | **1.28 (1.16, 1.41)** | **1.28 (1.14, 1.40)** | **1.28 (1.14, 1.40)** | **1.28 (1.16, 1.41)** |
| 5-9 | **1.40 (1.27, 1.53)** | **1.40 (1.27, 1.54)** | **1.40 (1.27, 1.54)** | **1.40 (1.27, 1.53)** |
| 10-14 | **1.36 (1.24, 1.48)** | **1.36 (1.25, 1.48)** | **1.36 (1.25, 1.48)** | **1.36 (1.24, 1.48)** |
| 15-19 | 0.96 (0.86, 1.07) | 0.96 (0.87, 1.06) | 0.96 (0.87, 1.06) | 0.96 (0.87, 1.06) |
| 20-24 | **0.74 (0.67, 0.82)** | **0.74 (0.66, 0.82)** | **0.74 (0.66, 0.82)** | **0.74 (0.67, 0.82)** |
| 25-34 | **0.74 (0.69, 0.80)** | **0.74 (0.68, 0.79)** | **0.74 (0.68, 0.79)** | **0.74 (0.69, 0.80)** |
| 35-44 | **0.88 (0.82, 0.94)** | **0.88 (0.82, 0.94)** | **0.88 (0.82, 0.94)** | **0.88 (0.82, 0.95)** |
| 45-54 | **0.86 (0.80, 0.91)** | **0.86 (0.81, 0.91)** | **0.86 (0.81, 0.91)** | **0.86 (0.81, 0.91)** |
| 55-64 | 0.95 (0.90, 1.01) | 0.95 (0.90, 1.01) | 0.95 (0.90, 1.01) | 0.95 (0.90, 1.01) |
| 65-69 | 0.98 (0.92, 1.04) | 0.98 (0.92, 1.04) | 0.98 (0.92, 1.04) | 0.98 (0.92, 1.05) |
| 70-74 | 1.06 (0.98, 1.14) | 1.06 (0.98, 1.15) | 1.06 (0.98, 1.15) | 1.06 (0.97, 1.14) |
| 75-79 | 0.93 (0.82, 1.05) | 0.93 (0.81, 1.05) | 0.93 (0.81, 1.05) | 0.93 (0.81, 1.05) |
| 80-85 | 1.07 (0.88, 1.26) | 1.07 (0.88, 1.25) | 1.07 (0.88, 1.25) | 1.06 (0.88, 1.27) |
| HH Size 1 | **1.20 (1.14, 1.25)** | **1.20 (1.14, 1.25)** | **1.20 (1.14, 1.25)** | **1.20 (1.14, 1.25)** |
| HH Size 2 | **1.06 (1.01, 1.10)** | **1.06 (1.01, 1.10)** | **1.06 (1.01, 1.10)** | **1.06 (1.01, 1.11)** |
| HH Size 3 | **0.94 (0.89, 0.98)** | **0.94 (0.90, 0.98)** | **0.94 (0.90, 0.98)** | **0.94 (0.90, 0.98)** |
| HH Size 4 | 0.98 (0.92, 1.02) | 0.98 (0.92, 1.02) | 0.98 (0.92, 1.02) | 0.98 (0.92, 1.03) |
| HH Size 5+ | **0.86 (0.79, 0.93)** | **0.86 (0.79, 0.93)** | **0.86 (0.79, 0.93)** | **0.86 (0.79, 0.93)** |
| Weekday | 0.97 (0.95, 1.01) | 0.97 (0.95, 1.01) | 0.97 (0.95, 1.01) | 0.97 (0.95, 1.01) |
| Weekend | 1.03 (0.99, 1.06) | 1.03 (0.99, 1.05) | 1.03 (0.99, 1.05) | 1.03 (0.99, 1.06) |
| Employed full-time | **1.37 (1.30, 1.44)** | **1.37 (1.30, 1.44)** | **1.37 (1.30, 1.44)** | **1.37 (1.30, 1.44)** |
| Employed part-time | **1.42 (1.34, 1.51)** | **1.42 (1.34, 1.51)** | **1.42 (1.34, 1.51)** | **1.42 (1.34, 1.51)** |
| Full-time parent | **0.55 (0.46, 0.65)** | **0.55 (0.46, 0.64)** | **0.55 (0.46, 0.64)** | **0.55 (0.46, 0.63)** |
| Long-term sick or disabled | 0.99 (0.87, 1.08) | 0.99 (0.87, 1.08) | 0.99 (0.87, 1.08) | 0.99 (0.87, 1.08) |
| Retired | 0.97 (0.90, 1.02) | 0.97 (0.90, 1.02) | 0.97 (0.90, 1.02) | 0.97 (0.91, 1.02) |
| Self-employed | 1.10 (0.99, 1.20) | 1.10 (0.99, 1.20) | 1.10 (0.99, 1.20) | 1.10 (0.99, 1.20) |
| Student (15+) | **1.16 (1.05, 1.27)** | **1.16 (1.06, 1.27)** | **1.16 (1.06, 1.27)** | **1.16 (1.07, 1.28)** |
| Unemployed, not seeking | 0.90 (0.78, 1.04) | 0.90 (0.77, 1.03) | 0.90 (0.77, 1.03) | 0.90 (0.77, 1.02) |
| Unemployed, seeking | **0.82 (0.71, 0.92)** | **0.82 (0.72, 0.93)** | **0.82 (0.72, 0.93)** | **0.82 (0.72, 0.93)** |
| Asymptomatic | **0.83 (0.80, 0.85)** | **0.83 (0.80, 0.85)** | **0.83 (0.80, 0.85)** | **0.83 (0.80, 0.85)** |
| Symptomatic | **1.20 (1.17, 1.23)** | **1.20 (1.18, 1.23)** | **1.20 (1.18, 1.23)** | **1.20 (1.18, 1.23)** |
| With mask | **1.42 (1.38, 1.47)** | **1.42 (1.38, 1.47)** | **1.42 (1.38, 1.47)** | **1.42 (1.38, 1.46)** |
| Without mask | **0.69 (0.66, 0.71)** | **0.69 (0.66, 0.71)** | **0.69 (0.66, 0.71)** | **0.69 (0.66, 0.71)** |
| Low concern about severity | **1.11 (1.07, 1.15)** | **1.11 (1.07, 1.15)** | **1.11 (1.07, 1.15)** | **1.11 (1.07, 1.15)** |
| Neutral on severity | 0.97 (0.93, 1.01) | 0.97 (0.93, 1.01) | 0.97 (0.93, 1.01) | 0.97 (0.93, 1.01) |
| Concern about severity | **0.93 (0.89, 0.96)** | **0.93 (0.90, 0.96)** | **0.93 (0.90, 0.96)** | **0.93 (0.90, 0.96)** |
| Low infection possibility | 1.04 (1.00, 1.08) | **1.04 (1.01, 1.08)** | **1.04 (1.01, 1.08)** | **1.04 (1.01, 1.08)** |
| Neutral on possibility | 1.00 (0.97, 1.03) | 1.00 (0.97, 1.03) | 1.00 (0.97, 1.03) | 1.00 (0.97, 1.03) |
| High infection possibility | 0.96 (0.92, 1.00) | 0.96 (0.92, 1.00) | 0.96 (0.92, 1.00) | 0.96 (0.92, 1.00) |
| Distrust gov advice | 1.00 (0.95, 1.03) | 1.00 (0.95, 1.03) | 1.00 (0.95, 1.03) | 1.00 (0.95, 1.03) |
| Neutral on gov advice | **0.89 (0.86, 0.93)** | **0.89 (0.86, 0.93)** | **0.89 (0.86, 0.93)** | **0.89 (0.86, 0.93)** |
| Trust gov advice | **1.12 (1.08, 1.16)** | **1.12 (1.08, 1.16)** | **1.12 (1.08, 1.16)** | **1.12 (1.08, 1.16)** |

Estimated relative risk (posterior median with 95% CrI) of having daily non-household contacts in the top 20% of the population within the given exposure group, compared to the population average. Bold values indicate statistical significance at the 95% credible level (i.e., the 95% CrI does not include 1).

### A.2 Data

#### A.2.1 The COVIMOD study

##### Data preprocessing

We excluded contacts for whom age or gender information was not provided, as our analysis relies on this demographic information. A total of 171 such contacts were excluded, accounting for 0.29% of the data. For ethical reasons, the ages of children under 18 years old were reported in age groups (0-4, 5-9, 10-14, and 15-19). We interpolated the ages of children using discrete uniform distributions, with the ranges corresponding to the reported age groups. For 175 participants aged 85 and older, we set their age to 85 in the analysis because there were too few data points at ages 85 and above for contact patterns to be analyzed at 1-year resolution (Supplementary Fig. 8). We identified one potentially anomalous report exceeding 2000 contacts and set it to the maximum reported value below this threshold during that period (*k* = 817).

#### A.2.2 Other datasets

Daily case and death data were retrieved from COVID-19 Data Repository by the Center for Systems Science and Engineering (CSSE) at Johns Hopkins University. ^1^ The map layers of Germany and surrounding areas were sourced from OpenStreetMap. ^2^ The county-level geographical boundaries were adjusted using shapefile from Opendatasoft ^3^ to match the granularity required by the COVIMOD study. The geographical division is determined by the Nomenclature of Territorial Units for Statistics (NUTS) ^4^ level three codes from the 2021 edition. Population size estimates by age and sex for 2021 were obtained from the GENESIS-online database ^5^ (table code: 12411-0041).

### A.3 Software

All analysis in this study were performed using Python 3.10. Bayesian inference used the No-U-turn sampler (NUTS) ^6^ provided by the *Stan* ^7^ probabilistic programming language via command-line interface `CmdStan` (version 2.35.0). Epidemic simulation were done using the stochastic agent-based simulator Covasim ^8^ (version 3.1.6). Analyses were conducted on the HPC at the Saw Swee Hock School of Public Health, National University of Singapore; the majority of results can be reproduced on a modern laptop.

### A.4 Statistical analysis of non-household contact distributions

#### A.4.1 The Beta-Negative Binomial (BNB) distribution

The BNB distribution is a family of discrete distributions with infinite support, ^9^ whose main properties have been studied by Irwin ^10–13^ and Xekalaki. ^14–18^ For *r >* 0, *α >* 0 and *β >* 0, for *y* ∈ ℕ ∪ {0}, the probability mass function (pmf) is given by

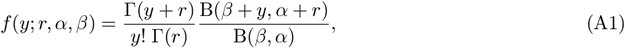

where Γ(*x*) and B(*x, y*) are Gamma and Beta functions, respectively. The BNB is scale-free in the sense that its right tail follows a power-law distribution. Specifically, as *y* → ∞,

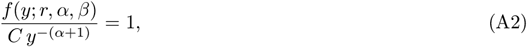

where *C* is a constant depending on the parameters. Based on Eq. (A2), the heavy right tail is characterised by the tail exponent, or so-called scale-free exponent *α* + 1.

In the probability mass function (pmf) of the BNB distribution, the parameters *r* and *β* are symmetric, leading to non-identifiability. We therefore propose a reparameterization: for *µ >* 0, *σ >* 0, and *γ >* 0, we defined

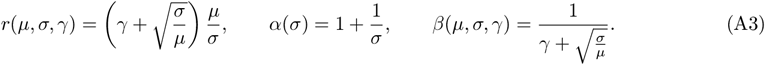

Then the pmf for the new parameterization is

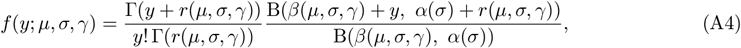

where *µ* represents the mean, *σ* controls tail heaviness, and *γ* is a dispersion parameter. Specifically,

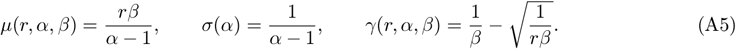

#### A.4.2 Comparison of heavy tail models

In order to identify the functional form that best describes the distribution of contact network degree data, we compared seven distributions: Poisson, Negative Binomial, Geometric, Zipf (discrete Pareto), Yule–Simon, Waring, and BNB. The first three distributions are widely used to model count data that are not characterized by heavy tails, and the Zipf, Yule–Simon, and Waring distributions can be used to capture heavy tails, for example for modelling sexual contact data. ^19–22^

Let *Y*_*i*_ denote the number of non-household contacts reported by the *i*th of *i* = 1, …, *n* respondents in each of the five phases shown in Figure 1. We modelled *Y*_*i*_ using the 7 distributions above, attaching linear predictors of the form

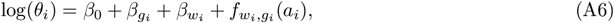

where *θ*_*i*_ denotes either mean *µ*_*i*_ or tail exponent *α*_*i*_ depending on the distribution. Additionally, *g*_*i*_ ∈ {*M, F* } denotes gender, *w*_*i*_ ∈ {1, …, 5} pandemic phase, and *a*_*i*_ ∈ [0, 85] age. Gender and pandemic phase were modelled with fixed effects, and age through non-parametric functions *f* for each gender and phase, with priors

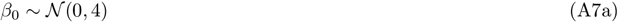

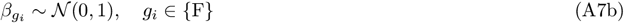

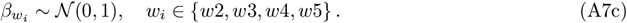

The interaction term 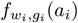 follows a Hilbert-Space Gaussian process prior ^23^

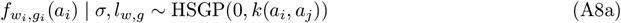

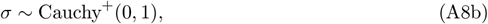

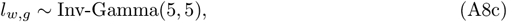

where *k*(*a*_*i*_, *a*_*j*_) represents a squared exponential kernel with variance *σ* and lengthscale *l*_*w,g*_. Priors on *σ* and *l*_*w,g*_ were specified along standard recommendations. ^24^

To compare models, we ran experiments on first-time participants (*n*=7,834), using as metric for comparison the expected log pointwise predictive density (ELPD) estimated with leave-one-out cross-validation (LOO-CV). ^25^ BNB significantly outperformed all other models for each wave (Supplementary Fig. 1 and Supplementary Table 1). The thin-tailed models (Poisson, negative binomial and geometric) did not pass the diagnostics for Pareto smoothed importance sampling, ^26^ indicating that these models were unstable in capturing the extremes in the data.

#### A.4.3 Bayesian inference framework

We built a Bayesian hierarchical model to estimate the non-household contact distribution by age, gender, and pandemic phase from all survey respondents regardless of repeat participation, *i* = 1, …, 59, 414. This model subsequently formed the basis for obtaining population-level estimates through post-stratification (Section A.4.4) as shown in Figure 1 in the main text.

Let *Y*_*i*_ denote the number of non-household contacts reported by the *i*th respondent, which we model with the BNB distribution described above. We attached linear predictors to all three model parameters, the component 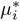 of the mean that is unrelated to the tail, tail heaviness *σ*_*i*_, and dispersion *γ*_*i*_:

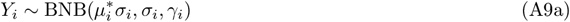

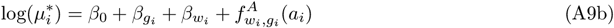

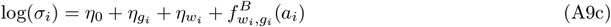

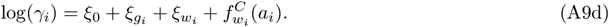

All parameters were assigned the following priors, and followed weakly informative, standard recommendations as available, ^24,27^

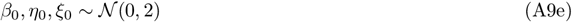

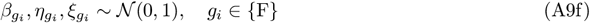

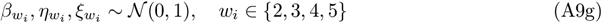

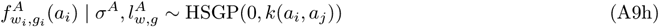

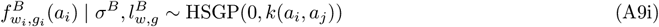

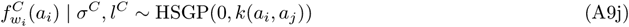

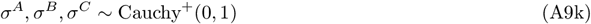

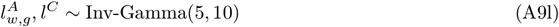

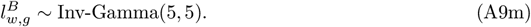

We ran four MCMC chains in parallel to assess convergence and mixing, with 1000 warm-up iterations and 1000 sampling iterations per chain. The samples did not exhibit any convergence issues (Supplementary Fig. 7).

To estimate geographic differences in contact distributions shown in Figure 3 in the main text, we added to each of the three linear predictors of our model Eqs. (A9) three fixed effects that represented the three NUTS regions of the classification proposed by Schuppert et al. ^28^ . We additionally included pandemic phase-region interaction terms, and made the non-parametric age effects depend on pandemic phase, gender, and region. The region-specific model was numerically fitted as the base model above, with no convergence issues.

#### A.4.4 Post-stratification

The age- and sex-distribution of COVIMOD survey participants did not match that of the German population (Supplementary Fig. 8). For this reason, we applied post-stratification weighting to obtain population-representative estimates of non-household contact patterns.

Similar to the construction of multilevel regression and post-stratification (MRP) estimators,^30–32^ the posterior population-level average of non-household contacts 𝔼 [*Y*_pop,*w*_] and the posterior probability of having *y* non-household contacts *f*_pop,*w*_(*y*) during each pandemic phase *w* can be estimated as

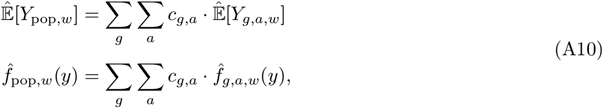

where the post-stratification weights *c*_*g,a*_ = *N*_*g,a*_*/N* were derived from the population size estimates by age and sex for 2021. ^5^

The tail exponent is not a linear statistical functional, and we employed a numerical post-stratification approach. Specifically, we drew *b* = 1, …, *B* samples from the joint posterior distribution *θ*^(*b*)^ ∼ *p*(*θ* | 𝒟). Next, for each Monte Carlo draw *b*, we generated a synthetic set of participants *i*^∗^ = 1, …, *n* of the same size as the original data, whose age and gender distribution matched that of the German population. Then, for each such synthetic participant *i*^∗^, we generated one observation 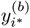 of non-household contacts from the posterior predictive distribution under model (A9). Thus, the predicted dataset 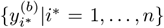 of non-household contacts corresponds to a synthetic set of participants of the same sample size as the original survey, that also matches the German population by age and gender. Finally, for each Monte Carlo draw *b*, we re-fitted model (A9) but since the synthetic sample matched age-and-sex characteristics of the entire population, we excluded all age–gender–related components. This yielded numerical estimates of the tail exponent 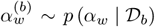. We pooled these and reported in Figs. 2-3 in the main text the median and equal-tailed 95% credible intervals across the pooled numerical samples.

**Supplementary Figure 7.**
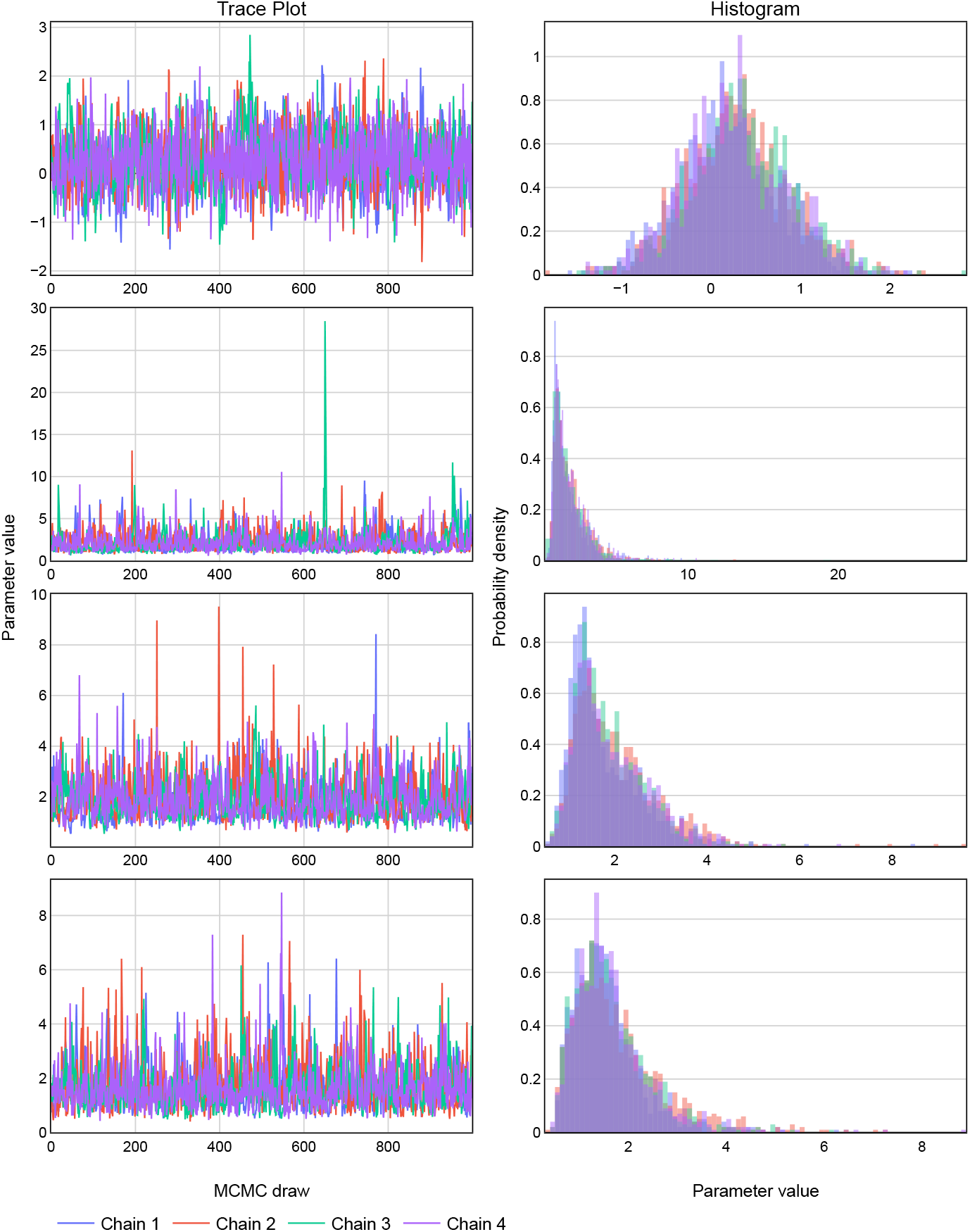
MCMC diagnostic plots for the four parameters with the highest R-hat statistic. Those R-hat statistics are from 1.00 to 1.01, and effective sample sizes ranged from 191 to 753. The left column shows the trace plots for four independent chains (indicated by different colors). The right column shows the corresponding posterior density. This plot across chains suggest good mixing and convergence to a common target distribution.

**Supplementary Figure 8.**
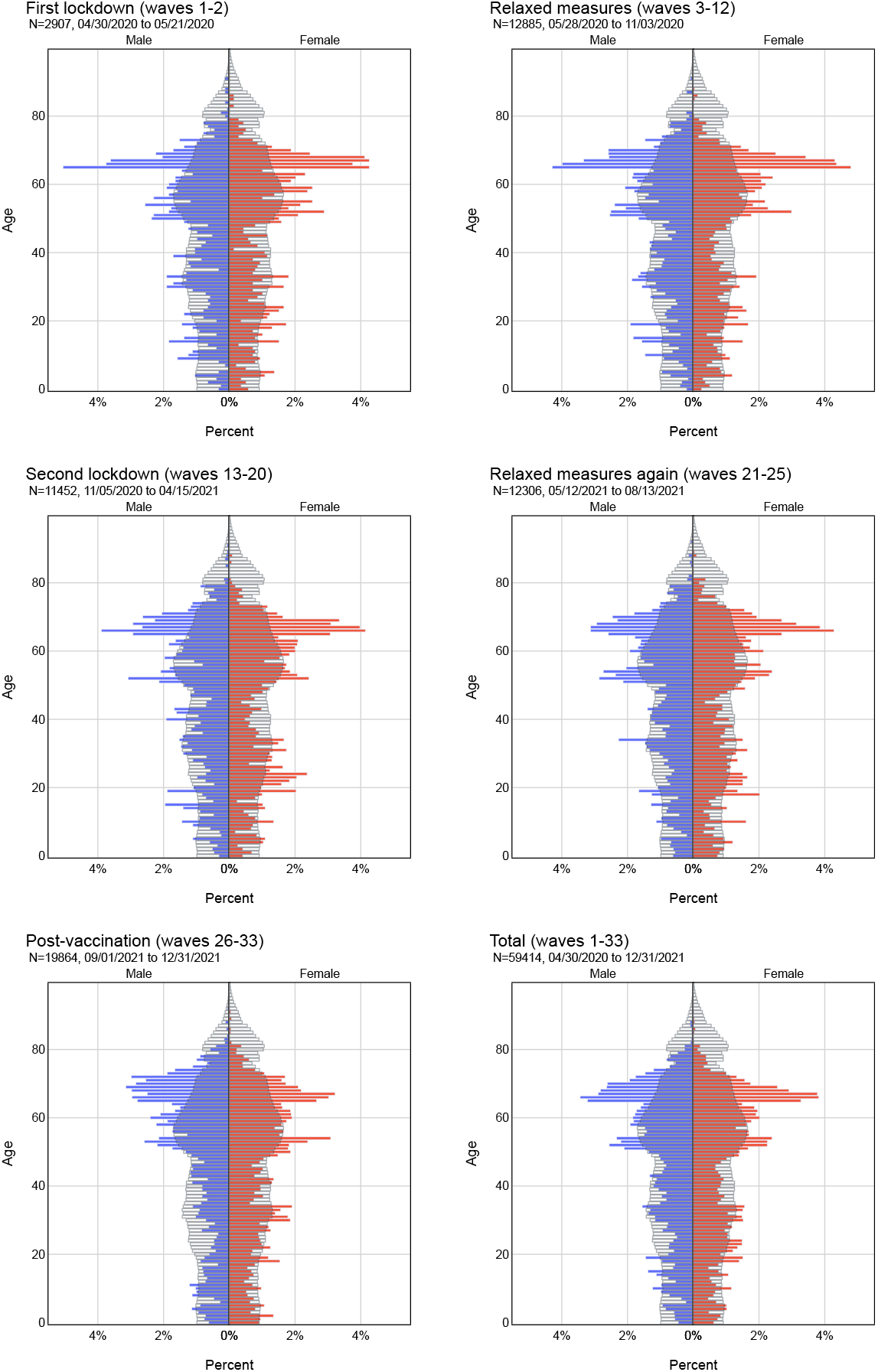
Age- and sex-specific distribution of COVIMOD survey participants versus the German population. The blue (red) bars represent the age distribution of male (female) participants, across the corresponding survey waves (panel). The underlying black bars show the estimated age-sex structure for 2021 as provided by the Federal Statistical Office of Germany. ^29^

#### A.4.5 Feature selection

We adopted a standard Bayesian variable selection framework to select for demographic and behavioural features associated with being in the tail. For each pandemic phase, we considered the upper-*p*-percentile *y*_*p*_ of the non-household contact distribution as the tail, with *p* ∈ {5%, 10%, 20%}, and then defined for each respondent a binary variable indicating whether their number of non-household contacts was in the tail, 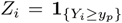. Here, for each phase, the upper-*p*-percentile was determined from the post-stratified, posterior predictive distribution of *Y*_*i*_ described in Section A.4.4, and therefore accounted for age- and sex-specific representation bias in the COVIMOD survey. This allowed us to conduct feature selection using a standard Bernoulli model,

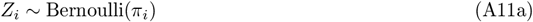

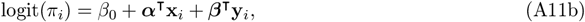

where x_*i*_ and y_*i*_ are aggregated control and test features, respectively,

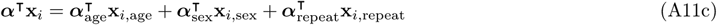

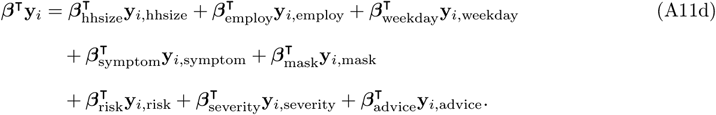

The control features allowed to account for known confounders without variable selection, while the test features were subjected to variable selection to identify those associated with being in the tail of the contact distribution. The test features included household size (1, 2, 3, 4, 5+), employment status (Employed, full-time; Employed, part-time; Full-time parent; Long-term sick/disabled; Retired; Self employed; Student (15+); Unemployed, seeking; Unemployed, not seeking), day of week (weekday vs weekend), presence of COVID-like symptoms (yes, no), whether a face mask was worn on the day of the survey (yes, no), risk perception (“I am likely to catch coronavirus”; agree, neutral, disagree), perceived severity (“Coronavirus would be a serious illness for me”; agree, neutral, disagree), and attitudes toward government advice (“If I don’t follow the government’s advice, I might spread coronavirus to someone who is vulnerable”; agree, neutral, disagree). We tested these features using sparsity-inducing regularised horseshoe priors. ^33^ Model (A11) was fitted to the non-household contact reports of *n* = 59, 414 respondents regardless of repeat participation with *Stan*, following the same inferential setup as before. Four MCMC chains were run in parallel, with 1000 warm-up iterations and 1000 sampling iterations per chain. We detected no convergence or mixing issues (Supplementary Fig. 9). The marginal posterior densities (Supplementary Fig. 10) illustrate the shrinkage effect of the regularised horseshoe prior while allowing strong signals to emerge.

Following numerical sampling from the joint posterior density, we approximated the posterior risk ratio (RR) of each control and test feature for being associated with the heavy tail,

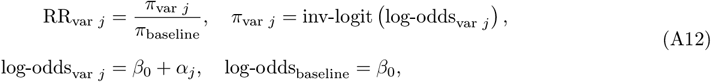

and report in Table 1 in the main text the median and 95% credible interval of the numerically approximated posterior risk ratios RR_*j*_.

### A.5 Modelling the impact of counterfactual interventions

#### A.5.1 Calibrated synthetic contact network

We initialized the population using the Hybrid network option in Covasim. ^8^ The simulation comprised 3.3 million agents, representing the German population at a 1:25 scale, with an age distribution in 10-year bands according to German census data. We used a standard 3-layer contact network including school (S), workplace and community (WC), and household layers (H). All children and adolescents aged 6 to 22 were assigned to schools and higher education institutions. Their number of non-household contacts in the S layer followed a Poisson distribution calibrated to reported daily effective contacts across 9 European countries,^34^ with a mean of 11.41. Adults aged 23 or over were assigned to the workplace and community layer, and their contact degree distribution in these layers was calibrated to our population-level estimates for Germany during the post vaccination period (September 2021–December 2021). The distribution for the number of daily contacts in the S and WC layers is shown in Fig. 12. Each person was randomly assigned to a household with the household size distribution matching the 2020 household projection^5^ (GENESIS-online database, table code: 12421-0100). We set the size of all households recorded as “5 or more” to 5. Supplementary Fig. 11 shows a comparison of the synthetic population and census data by age and household size.

#### A.5.2 Agent-based epidemic simulations

We used a stochastic discrete-time compartmental model to simulate the transmission dynamics of three template pathogens in an entirely susceptible population: SARS-CoV-2 wild strain, SARS-CoV-2 Omicron strain, and influenza A (H3N2). The model included five disease states: susceptible, exposed, infected, recovered, and deceased individuals. Infected individuals were further classified by symptom status (asymptomatic, pre-symptomatic, mild disease, severe disease, or critical disease), following the disease progression framework in Fig. 1 of Kerr et al. ^8^ . Table 1 of Kerr et al. ^8^ describes disease progression parameters for the SARS-CoV-2 wild strain, and Supplementary Tables 4-5 present the parameter values that we used for epidemic simulations of the SARS-CoV-2 Omicron and influenza A (H3N2) template pathogens. Supplementary Table 6 lists the values of all other parameters that we used for our epidemic simulation on the synthetic contact network, along with the primary literature reference. From the specified *R*_0_, we calibrated the transmission probability *β* per contact (edge) per day for each pathogen based on the degree distribution of the synthetic contact network during the post-vaccination period according to

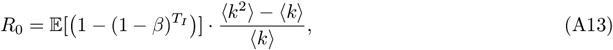

where ⟨*k*⟩ and ⟨*k*^2^⟩ are the first and second moments of the degree distribution of the German non-household contact network during the post-vaccination phase, and *T*_*I*_ is the infectious period. ^35^ In Eq. (A13), we approximated the transmissibility term 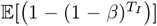 with 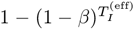, where 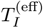 is the mean effective infectious period and itself approximated by

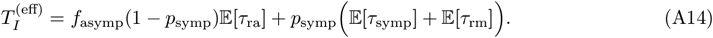

**Supplementary Figure 9.**
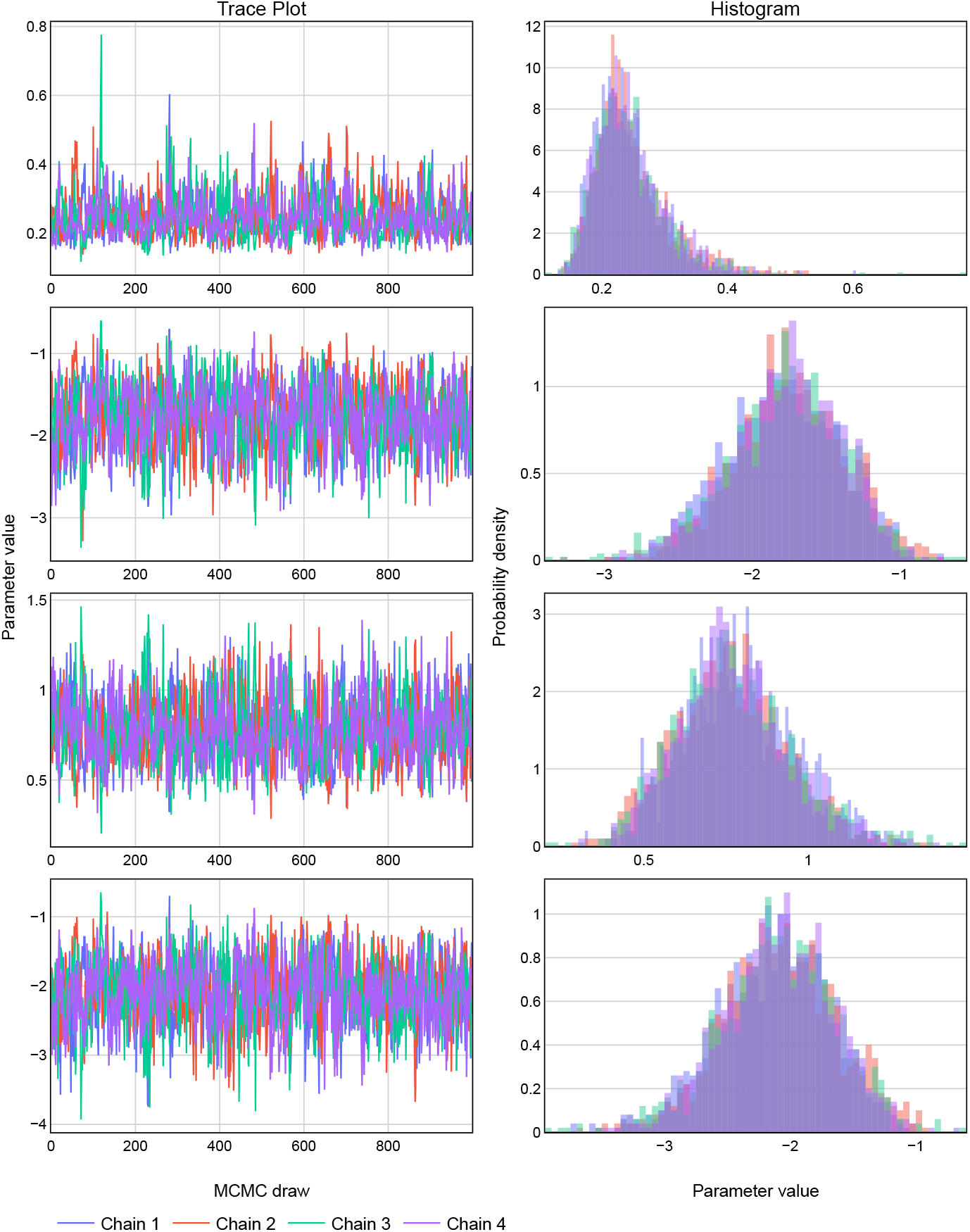
MCMC diagnostic plots for the four parameters with the lowest effective sample size in the 20% tail model. The effective sample sizes ranged from 757 to 924. The left column shows the trace plots for four independent chains (indicated by different colors). The right column shows the corresponding posterior densities.

**Supplementary Figure 10.**
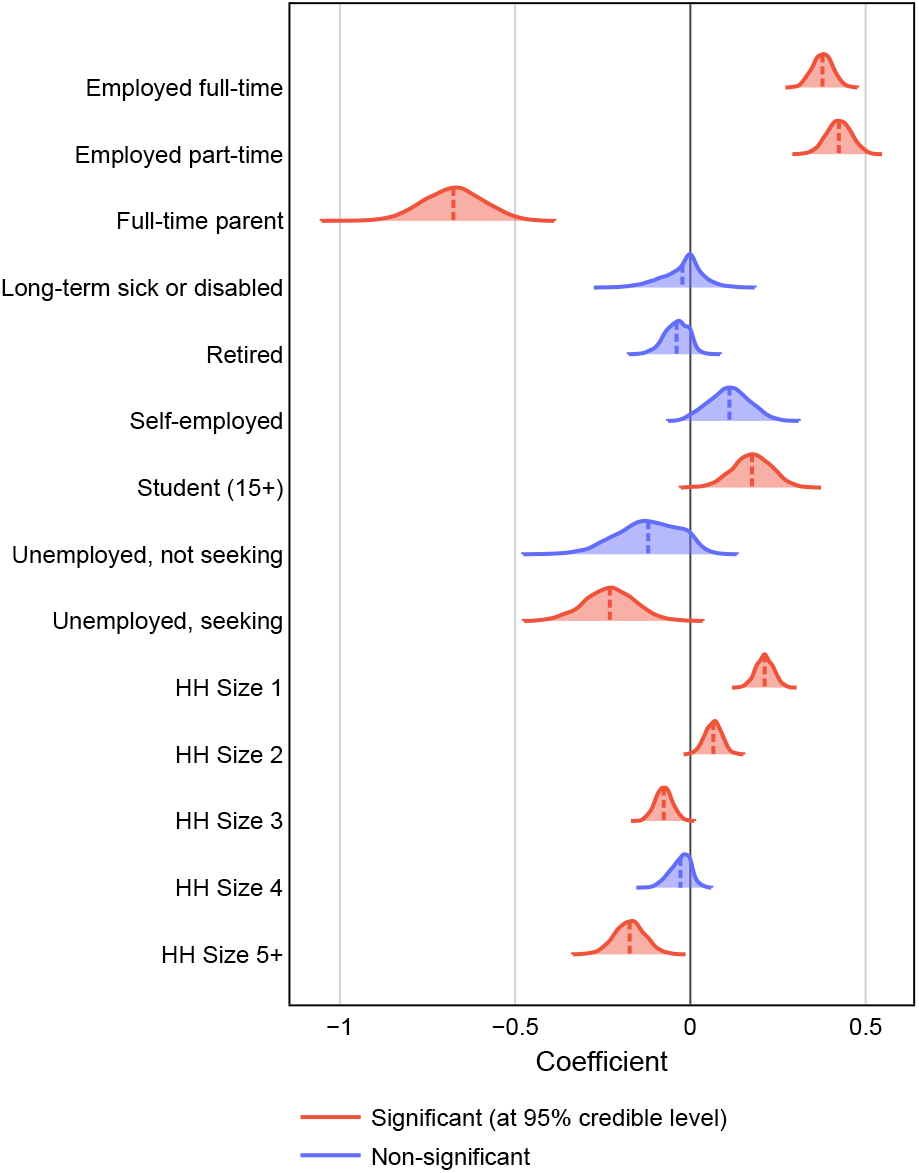
Marginal posterior densities of employment status and household size coefficients in the 20% tail model. Significant features (in red) are variables whose 95% highest density interval (HDI) does not contain zero. Non-significant features (in blue) exhibit densities centered around or overlapping zero.

**Supplementary Figure 11.**
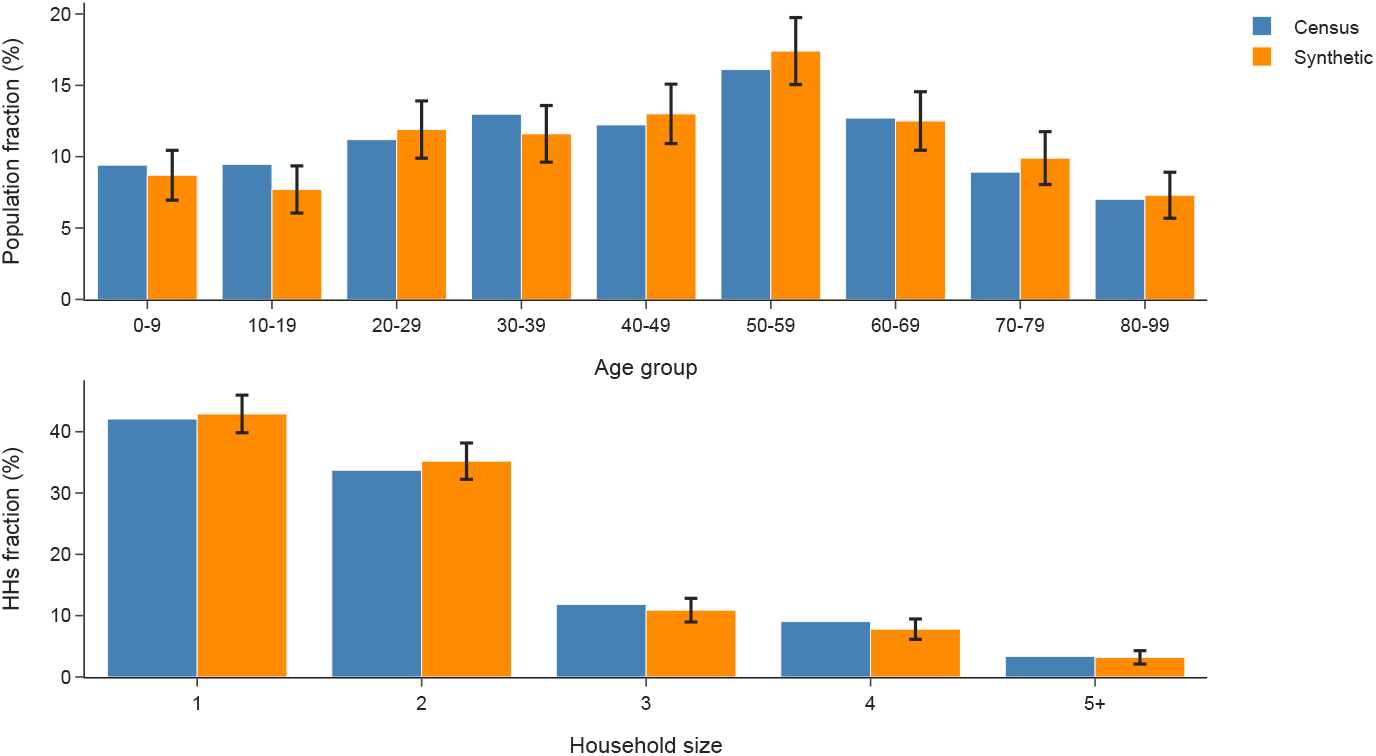
Age and household size distribution for the synthetic population versus the German census data. In each simulation, the agents and household structures are randomly initialized based on the German population features (blue bars). The error bars in the figure represent the 95% confidence intervals of the synthetic population’s relevant characteristics across 1,000 simulations.

**Supplementary Figure 12.**
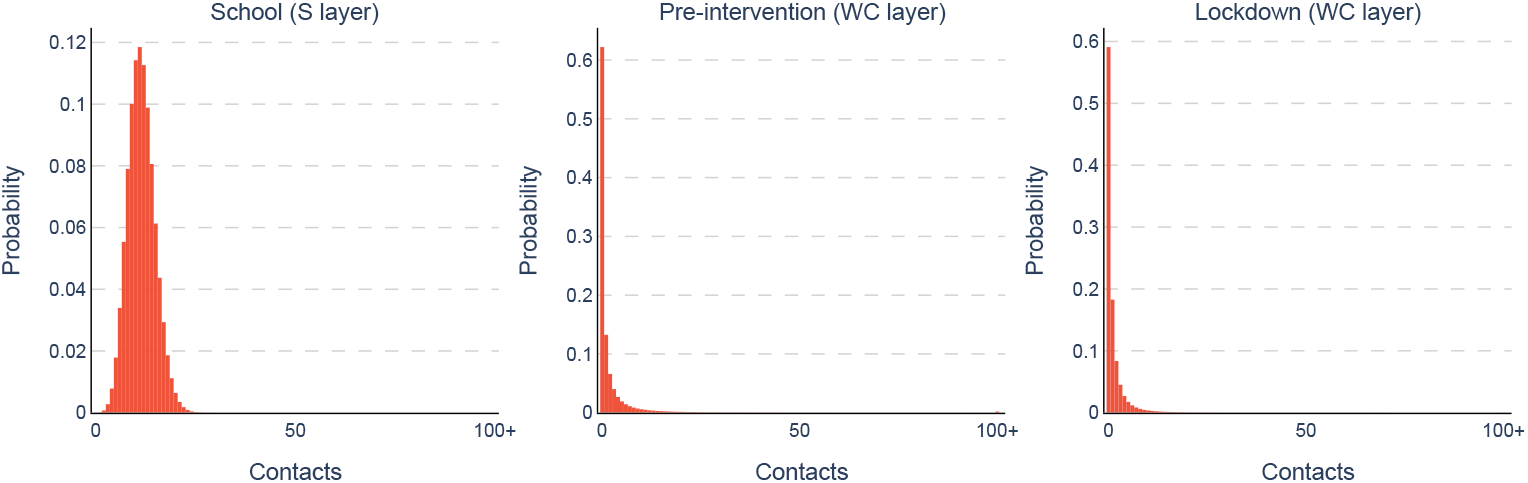
Degree distributions of the S and WC layers. Left: Degree distribution in the S layer for agents aged 6–22. Middle and Right: Degree distributions in the WC layer for adults (aged 23+) during the pre-intervention phase and under lockdown, respectively.

In Eq. (A14), we neglected the severe and critical branches because they constitute a very small fraction of cases in our parameterization. This allowed us to solve for *β* using the first-order approximation

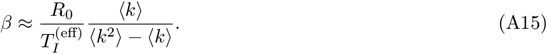

**Supplementary Table 4.** Disease progression parameterization for a SARS-CoV-2 Omicron (BA.1/BA.2) variant in epidemic simulations on the synthetic contact network.

| Parameter | Description | Distribution<br>(mean,std) | Ref. |
| --- | --- | --- | --- |
| $\tau_{\text{inf}}$ | Exposed-to-infectious | lognormal(3.1, 1.0) | [36] |
| $\tau_{\text{symp}}$ | Infectious-to-symptomatic | lognormal(0.7, 0.5) | [36] |
| $\tau_{\text{sev}}$ | Symptomatic-to-severe | lognormal(3.0, 2.0) | [37] |
| $\tau_{\text{cri}}$ | Severe-to-critical | lognormal(1.5, 2.0) | [8] |
| $\tau_{\text{dea}}$ | Critical-to-death | lognormal(10.7, 4.8) | [8] |
| $\tau_{\text{ra}}$ | Time from infectiousness onset to recovery for asymptomatic cases | lognormal(7.0, 2.0) | [38] |
| $\tau_{\text{rm}}$ | Time from symptom onset to recovery for mild cases | lognormal(6.9, 1.5) | [39] |
| $\tau_{\text{rs}}$ | Time from onset of severe symptoms to recovery for severe cases | lognormal(6.0, 2.0) | [40] |
| $\tau_{\text{rc}}$ | Time from onset of critical symptoms to recovery for critical cases | lognormal(5.0, 3.0) | [41] |

**Supplementary Table 5.** Disease progression parameterization for a influenza A (H3N2) variant in epidemic simulations on the synthetic contact network.

| Parameter | Description | Distribution<br>(mean,std) | Ref. |
| --- | --- | --- | --- |
| $\tau_{\text{inf}}$ | Exposed-to-infectious | lognormal(1.0, 0.5) | [42,43] |
| $\tau_{\text{symp}}$ | Infectious-to-symptomatic | lognormal(1.0, 0.5) | [42,43] |
| $\tau_{\text{sev}}$ | Symptomatic-to-severe | lognormal(3.0, 2.0) | [44] |
| $\tau_{\text{cri}}$ | Severe-to-critical | lognormal(2.0, 1.5) | [44,45] |
| $\tau_{\text{dea}}$ | Critical-to-death | lognormal(7.0, 4.0) | [46] |
| $\tau_{\text{ra}}$ | Time from infectiousness onset to recovery for asymptomatic cases | lognormal(5.0, 2.0) | [47] |
| $\tau_{\text{rm}}$ | Time from symptom onset to recovery for mild cases | lognormal(5.0, 2.0) | [48,49] |
| $\tau_{\text{rs}}$ | Time from onset of severe symptoms to recovery for severe cases | lognormal(8.0, 3.0) | [50] |
| $\tau_{\text{rc}}$ | Time from onset of critical symptoms to recovery for critical cases | lognormal(8.0, 4.0) | [46] |

**Supplementary Table 6.** Further disease transmission parameters used in epidemic simulations on the synthetic contact network.

| Parameter | Description | Value | Ref. |
| --- | --- | --- | --- |
| SARS-CoV-2 wild strain |  |  |  |
| $R_0$ | basic reproduction number | 2.8 | [51] |
| $p_{\text{symp}}$ | prob. of developing symptoms | † | [52] |
| $p_{\text{sev}}$ | prob. of developing severe symptoms | † | [53] |
| $p_{\text{cri}}$ | prob. of developing critical symptoms | † | [53] |
| $p_{\text{dea}}$ | prob. of death | † | [54] |
| $f_{\text{asympt}}$ | relative infectiousness of asymptomatic carriers | 1.0 | [55] |
| $(m_h, m_s, m_{wc})$ | multiplier of $\beta$ for household ($h$), school ($s$), and workplace & community ($wc$) layer | (3.0, 0.6, 0.6) | [8] |
| SARS-CoV-2 Omicron |  |  |  |
| $R_0$ | basic reproduction number | 9.5 | [56] |
| $p_{\text{symp,rel}}$ | multiplier for prob. of developing symptoms | 1.0 | [57] |
| $p_{\text{sev,rel}}$ | multiplier for prob. of severe given symptoms | 0.5 | [58] |
| $p_{\text{cri,rel}}$ | multiplier for prob. of critical given severe | 0.6 | * |
| $p_{\text{dea,rel}}$ | multiplier for prob. of death given critical | 1.0 | * |
| $f_{\text{asympt}}$ | relative infectiousness of asymptomatic carriers | 1.0 | [55] |
| $(m_h, m_s, m_{wc})$ | multiplier of $\beta$ for household ($h$), school ($s$), and workplace & community ($wc$) layer | (3.0, 0.6, 0.6) | [8] |
| Influenza A (H3N2) |  |  |  |
| $R_0$ | basic reproduction number | 1.5 | [59] |
| $p_{\text{symp,rel}}$ | prob. of developing symptoms | 0.9 § | [60] |
| $p_{\text{sev,rel}}$ | prob. of developing severe symptoms | 4.6% § | * |
| $p_{\text{cri,rel}}$ | prob. of critical given severe | 0.17 | * |
| $p_{\text{dea,rel}}$ | prob. of death | 0.12% § | [61] |
| $f_{\text{asympt}}$ | relative infectiousness of asymptomatic carriers | 0.57 | [62] |
| $(m_h, m_s, m_{wc})$ | multiplier of $\beta$ for household ($h$), school ($s$), and workplace & community ($wc$) layer | (3.0, 2.0, 0.2) | * |

†: see Table 2 of Kerr et al. <sup>8</sup>; §: the average across all ages; age-specific values are actually used; \*: assumed.

#### A.5.3 Counterfactual interventions

We implemented the following synthetic counterfactual intervention strategies on each epidemic simulation.

##### Tailored protection scenario

At each time step of the epidemic simulation, remove at random from all non-household contacts those that link to the 5% of adults who have the most WC layer links.

##### School closure

After the number of nationwide deaths reaches 500^1^, remove all S layer links at each time step of the epidemic simulation.

##### Lockdown

After the number of nationwide deaths reaches 500, reduce the number of non-household contacts for all adults by resampling their degree in the WC layer from the estimated degree distribution during the first lockdown period in Germany. After this resampling step, synthesize a new WC layer network using the configuration model. Use this network at each following time step of the epidemic simulation.

##### Lockdown with school closure

After the number of nationwide deaths reaches 500, implement both lockdown and school closure.

##### Bubble 1 + 1

After the number of nationwide deaths reaches 500, implement both lockdown and school closure. Ease this intervention scenario by merging single-person households into 2-person households (a 2-bubble). Use this network at each following time step of the epidemic simulation. We assumed that the transmission rate for contacts between households within the same bubble is 50% lower than that for contacts within households.

##### Bubble 1 + n

After the number of nationwide deaths reaches 500, implement both lockdown and school closure. Ease this intervention scenario by merging single-person households with another household of arbitrary size (*n*=1,2,3,4,5) to form a bubble of size *n* + 1. Use this network at each following time step of the epidemic simulation. We assumed that the transmission rate for contacts between households within the same bubble is 50% lower than that for contacts within households.

##### Testing

Using the Robert Koch Institut report of daily COVID-19 tests per 1,000 people in Germany, ^63^ a specified number of tests were conducted daily at each time step of the epidemic simulation. Individuals who test positive were required to isolate at home, and we assumed full compliance. The test sensitivity was set to 0.97. ^64^ We followed the default setting in Covasim, in which individuals who developed COVID-19 symptoms were 100 times more likely to be tested than asymptomatic or uninfected individuals, to reflect that symptomatic individuals were prioritized for testing.

##### Contact tracing

When counterfactual testing was in effect, if an agent tested positive, a proportion of their contacts were immediately notified of disease exposure (100% of household contacts, 50% of school contacts and 30% of workplace and community contacts). After notification, all agents complied and self-isolated for the following 14 days in the epidemic simulation.

For each pathogen and counterfactual scenario combination, we independently generated 1,000 synthetic populations (each comprising 3.3 million agents) with their contact networks. Then one epidemic simulation was run on each population, with 0.1% of agents randomly selected as initially infected individuals. Fig. 5 reports boxplots summarizing the distribution of infections and deaths at day 150 across these 1,000 independent simulations.

### A.6 Sensitivity analyses

#### A.6.1 Sensitivity to parental reporting of contacts on behalf of their children

In the COVIMOD survey, parents or guardians reported contacts on behalf of their dependent children (proxy-reporting) and this may have led to an underestimation of their number of contacts. To test the sensitivity of our results to this potential bias, we conducted analyses that excluded all proxy-reported entries that parents reported on behalf of their children, leaving us with data from *n*=49,933 respondents. Supplementary Fig. 5 compares the strength of the heavy tail when excluding proxy-reported entries to those in the main analysis, indicating that the 80/20 rule and the classification of phases into finite and infinite-variance regimes were robust to proxy reporting. We further explored the strength of association of demographic and behavioural features with the heavy tail after excluding proxy-reported contacts, focussing on the top 20% heavy tail in Supplementary Table 2. In the table, we show the posterior probabilities of significance for each feature, *p* = max{*p*_+_, *p*_−_}, where *p*_+_ = ℙ (RR_*j*_ *>* 1 | *D*), and *p*_−_ = ℙ (RR_*j*_ *<* 1 | *D*). We use ℙ (· | *D*) to denote the posterior probability. *p*_+_ and *p*_−_ were estimated by Monte Carlo integration. We found that most of the variables identified as significant in the main analysis remained significant (living alone; employed; full-time parent; symptomatic; using mask; etc.), while the posterior significance probability of other variables decreased only slightly.

#### A.6.2 Sensitivity to repeat participation

COVIMOD was a longitudinal study, and repeat participation in the survey has been associated with incomplete reporting of contacts, known as reporting fatigue. ^65^ In a sensitivity analysis, we restricted the data to first-time participants only (*n*=7,834) to assess the impact of reporting fatigue on our results. Supplementary Fig. 4 compares the strength of the heavy tail when restricting to first-time participants only to those in the main analysis. Due to the significantly reduced sample size, the uncertainty on tail inferences increased significantly. However, most of the posterior mass of the tail exponent remains between 2 and 3 for every phase after the first lockdown, in agreement with the findings in the main analysis. Considering the demographic and behavioural features associated with the heavy tail, we found that the large majority of the same features as in the main analysis remained associated with the top 20% heavy tail (Supplementary Table 2).

**Supplementary Table 7.**
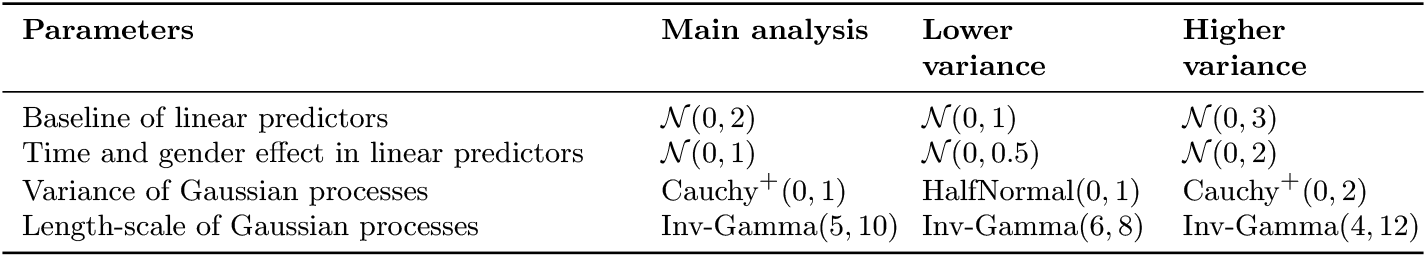
Specification of prior distributions on effect sizes in sensitivity analyses.

#### A.6.3 Sensitivity to the strength of prior distributions on effect sizes

We followed standard recommendations on specifying the prior distributions of all parameters of our main model Eq. (A9), ^24^ including the Inv-Gamma(5, 10) prior for GP length-scales, ^27^ however these choices could still influence our inferences. Therefore, in two sensitivity analyses, we constructed two alternative sets of prior settings for the regression coefficients and the GP variance and length-scale parameters, with the first comprising prior distributions of lower variance and the second comprising prior distributions of higher variance compared to those in the main analysis. Supplementary Table 7 summarizes these prior specifications. We report in Supplementary Fig. 6 the estimated population-level CCDF of non-household contacts across all pandemic phases under these two alternative prior settings. Across the prior settings explored, the model yielded consistent estimates that aligned well with the empirical data.

#### A.6.4 Sensitivity to the strength of prior distributions on feature selection

We also tested the sensitivity in the strength of association of behavioural and demographic features with the top 20% heavy tail under alternative prior specifications of our feature selection model. In the main analysis, we used sparsity-inducing priors with strong local shrinkage and a wide slab to avoid false discoveries. Here, we constructed two alternative prior settings: one with weaker local shrinkage and another with a narrower or wider slab. Specifically, in the main analysis we used the sparsity-inducing priors

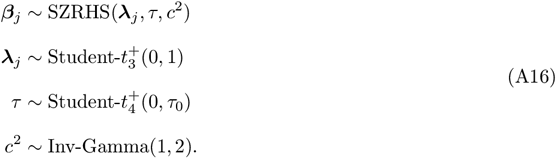

In practice, the sum-to-zero regularized horsehose prior (SZRHS) is very sensitive to the prior specification of the global shrinkage parameter *τ* . Its scale *τ*_0_ is determined by *p*_0_. ^66^ In the main analysis, we set the number of effective non-zero coefficients a priori to the theoretical minimum (i.e. set *p*_0_ = 1), corresponding to the strongest global shrinkage; a conservative choice in terms of variable selection. Thus, any selected signals were strongly supported by the likelihood rather than by the prior. In sensitivity analyses, we investigated alternative prior specifications on λ and *c*^2^. In the weaker local shrinkage analysis, we set

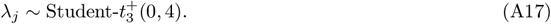

In a narrower slab analysis, we set

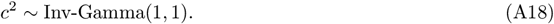

In a wider slab analysis, we set

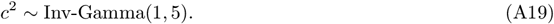

Supplementary Table 3 reports the strength of association of the behavioural and demographic features with the top 20% under these sensitivity assumptions, indicating that the posterior support for the associations reported in the main analysis remained essentially unchanged.

## Footnotes

1 The synthetic population represents the German population at a ratio of 1:25, so in the simulation this translates to 25 agent deaths

## Notes

### Competing Interest Statement

The authors have declared no competing interest.

### Funding Statement

This study was funded by National Medical Research Council, Singapore Ministry of Health (PREPARE-S2-2024-002), EPSRC Centre for Doctoral Training in Modern Statistics and Statistical Machine Learning at Imperial and Oxford (EP/S023151/1 to Prof. Axel Gandy), and National Research Foundation, Singapore (NRF-NRFF15-2023-0010).

### Author Declarations

The ethics committee of the Medical Board Westfalen-Lippe and the University of Munster gave ethical approval for this work (reference number 2020-473-fs).

## References

[1] Lloyd-Smith, J.O., Schreiber, S.J., Kopp, P.E., Getz, W.M.: Superspreading and the effect of individual variation on disease emergence. Nature 438(7066), 355–359 (2005) 10.1038/nature04153

[2] Fraser, C., Riley, S., Anderson, R.M., Ferguson, N.M.: Factors that make an infectious disease outbreak controllable. Proceedings of the National Academy of Sciences 101(16), 6146–6151 (2004) 10.1073/pnas.0307506101

[3] Haug, N., Geyrhofer, L., Londei, A., Dervic, E., Desvars-Larrive, A., Loreto, V., Pinior, B., Thurner, S., Klimek, P.: Ranking the effectiveness of worldwide COVID-19 government interventions. Nature Human Behaviour 4(12), 1303–1312 (2020) 10.1038/s41562-020-01009-0

[4] Flaxman, S., Mishra, S., Gandy, A., Unwin, H.J.T., Mellan, T.A., Coupland, H., Whittaker, C., Zhu, H., Berah, T., Eaton, J.W., et al.: Estimating the effects of non-pharmaceutical interventions on covid-19 in europe. Nature 584(7820), 257–261 (2020) 10.1038/s41586-020-2405-7

[5] Kraemer, M.U.G., Yang, C.-H., Gutierrez, B., Wu, C.-H., Klein, B., Pigott, D.M., Open COVID-19 Data Working Group, Du Plessis, L., Faria, N.R., Li, R., Hanage, W.P., Brownstein, J.S., Layan, M., Vespignani, A., Tian, H., Dye, C., Pybus, O.G., Scarpino, S.V.: The effect of human mobility and control measures on the COVID-19 epidemic in China. Science 368(6490), 493–497 (2020) 10.1126/science.abb4218

[6] Adam, D.C., Wu, P., Wong, J.Y., Lau, E.H.Y., Tsang, T.K., Cauchemez, S., Leung, G.M., Cowling, B.J.: Clustering and superspreading potential of SARS-CoV-2 infections in Hong Kong. Nature Medicine 26(11), 1714–1719 (2020) 10.1038/s41591-020-1092-0

[7] Pastor-Satorras, R., Vespignani, A.: Epidemic spreading in scale-free networks. Physical Review Letters 86(14), 3200 (2001)

[8] May, R.M., Lloyd, A.L.: Infection dynamics on scale-free networks. Physical Review E 64(6), (2001) 10.1103/PhysRevE.64.06611211736241

[9] Meyers, L.: Contact network epidemiology: Bond percolation applied to infectious disease prediction and control. Bulletin of the American Mathematical Society 44(1), 63–86 (2007) 10.1090/S0273-0979-06-01148-7

[10] Salathé, M., Kazandjieva, M., Lee, J.W., Levis, P., Feldman, M.W., Jones, J.H.: A high-resolution human contact network for infectious disease transmission. Proceedings of the National Academy of Sciences 107(51), 22020–22025 (2010) 10.1073/pnas.1009094108

[11] Abueg, M., Hinch, R., Wu, N., Liu, L., Probert, W., Wu, A., Eastham, P., Shafi, Y., Rosencrantz, M., Dikovsky, M., Cheng, Z., Nurtay, A., Abeler-Dörner, L., Bonsall, D., McConnell, M.V., O’Banion, S., Fraser, C.: Modeling the effect of exposure notification and non-pharmaceutical interventions on COVID-19 transmission in Washington state. npj Digital Medicine 4(49), 49 (2021) 10.1038/s41746-021-00422-7

[12] Isella, L., Romano, M., Barrat, A., Cattuto, C., Colizza, V., Broeck, W., Gesualdo, F., Pandolfi, E., Ravà, L., Rizzo, C., Tozzi, A.E.: Close encounters in a pediatric ward: measuring face-to-face proximity and mixing patterns with wearable sensors. PLoS ONE 6(2), 17144 (2011) 10.1371/journal.pone.001714421386902

[13] Fournet, J., Barrat, A.: Contact Patterns among High School Students. PLoS ONE 9(9), 107878 (2014) 10.1371/journal.pone.0107878

[14] Bailey, M., Cao, R., Kuchler, T., Stroebel, J., Wong, A.: Social Connectedness: Measurement, Determinants, and Effects. Journal of Economic Perspectives 32(3), 259–80 (2018) 10.1257/jep.32.3.259

[15] Zhang, J., Litvinova, M., Liang, Y., Wang, Y., Wang, W., Zhao, S., Wu, Q., Merler, S., Viboud, C., Vespignani, A., Ajelli, M., Yu, H.: Changes in contact patterns shape the dynamics of the COVID-19 outbreak in China. Science 368(6498), 1481–1486 (2020) 10.1126/science.abb8001

[16] Schlosser, F., Maier, B.F., Jack, O., Hinrichs, D., Zachariae, A., Brockmann, D.: COVID-19 lockdown induces disease-mitigating structural changes in mobility networks. Proceedings of the National Academy of Sciences 117(52), 32883–32890 (2020) 10.1073/pnas.2012326117

[17] Du, Y., Aoki, T., Fujiwara, N.: A review of human mobility: Linking data, models, and real-world applications. Journal of Computational Social Science 8(4), 90 (2025)

[18] Mossong, J., Hens, N., Jit, M., Beutels, P., Auranen, K., Mikolajczyk, R., Massari, M., Salmaso, S., Tomba, G.S., Wallinga, J., Heijne, J., Sadkowska-Todys, M., Rosinska, M., Edmunds, W.J.: Social contacts and mixing patterns relevant to the spread of infectious diseases. PLoS Medicine 5(3), 74 (2008) 10.1371/journal.pmed.0050074

[19] Jarvis, C.I.,, Zandvoort, K.V., Gimma, A., Prem, K., Klepac, P., Rubin, G.J., Edmunds, W.J.: Quantifying the impact of physical distance measures on the transmission of COVID-19 in the UK. BMC Medicine 18(1) (2020) 10.1186/s12916-020-01597-8

[20] Liu, C.Y., Berlin, J., Kiti, M.C., Fava, E.D., Grow, A., Zagheni, E., Melegaro, A., Jenness, S.M., Omer, S.B., Lopman, B., Nelson, K.: Rapid review of social contact patterns during the COVID-19 pandemic. Epidemiology 32(6), 781–791 (2021) 10.1097/ede.0000000000001412

[21] Verelst, F., Hermans, L., Vercruysse, S., Gimma, A., Coletti, P., Backer, J.A., Wong, K.L.M., Wambua, J., Zandvoort, K., Willem, L., Bogaardt, L., Faes, C., Jarvis, C.I., Wallinga, J., Edmunds, W.J., Beutels, P., Hens, N.: SOCRATES-CoMix: a platform for timely and open-source contact mixing data during and in between COVID-19 surges and interventions in over 20 european countries. BMC Medicine 19(1) (2021) 10.1186/s12916-021-02133-y

[22] Tomori, D.V., Rübsamen, N., Berger, T., Scholz, S., Walde, J., Wittenberg, I., Lange, B., Kuhlmann, A., Horn, J., Mikolajczyk, R., Jaeger, V.K., Karch, A.: Individual social contact data and population mobility data as early markers of SARS-CoV-2 transmission dynamics during the first wave in germany–an analysis based on the COVIMOD study. BMC Medicine 19(1) (2021) 10.1186/s12916-021-02139-6

[10] Irwin, J.O.: The generalized waring distribution applied to accident theory. Journal of the Royal Statistical Society Series A 131(2), 205 (1968) 10.2307/2343842

[24] Walde, J., Chaturvedi, M., Berger, T., Bartz, A., Killewald, R., Tomori, D.V., Rübsamen, N., Lange, B., Scholz, S., Treskova, M., et al.: Effect of risk status for severe covid-19 on individual contact behaviour during the sars-cov-2 pandemic in 2020/2021—an analysis based on the german covimod study. BMC Infectious Diseases 23(1), 1–11 (2023) 10.1186/s12879-023-08175-2

[26] Vehtari, A., Simpson, D., Gelman, A., Yao, Y., Gabry, J.: Pareto Smoothed Importance Sampling. arXiv (2024). 10.48550/arXiv.1507.02646

[26] Dorogovtsev, S.N., Mendes, J.F.F., Samukhin, A.N.: Structure of Growing Networks with Preferential Linking. Physical Review Letters 85(21), 4633–4636 (2000) 10.1103/PhysRevLett.85.4633

[27] Danon, L., House, T.A., Read, J.M., Keeling, M.J.: Social encounter networks: Collective properties and disease transmission. Journal of the Royal Society Interface 9(76), 2826–2833 (2012) 10.1098/rsif.2012.0357

[28] Read, J.M., Lessler, J., Riley, S., Wang, S., Tan, L.J., Kwok, K.O., Guan, Y., Jiang, C.Q., Cummings, D.A.T.: Social mixing patterns in rural and urban areas of southern China. Proceedings of the Royal Society B: Biological Sciences 281(1785), 20140268 (2014) 10.1098/rspb.2014.0268

[29] Clauset, A., Shalizi, C.R., Newman, M.E.: Power-law distributions in empirical data. SIAM Review 51(4), 661–703 (2009)

[30] Volz, E.: Sir dynamics in random networks with heterogeneous connectivity. Journal of Mathematical Biology 56(3), 293–310 (2008)

[31] Danon, L., Read, J.M., House, T.A., Vernon, M.C., Keeling, M.J.: Social encounter networks: Characterizing Great Britain. Proceedings of the Royal Society B: Biological Sciences 280(1765), 20131037 (2013) 10.1098/rspb.2013.1037

[32] Millest, A., Saeed, S., Symons, C., Carter, H.: Effect of face-covering use on adherence to other covid-19 protective behaviours: A systematic review. PLoS ONE 19(4), 0284629 (2024)

[33] Wu, Y., Song, S., Kao, Q., Kong, Q., Sun, Z., Wang, B.: Risk of sars-cov-2 infection among contacts of individuals with covid-19 in hangzhou, china. Public Health 185, 57–59 (2020)

[34] Ferretti, L., Wymant, C., Petrie, J., Tsallis, D., Kendall, M., Ledda, A., Di Lauro, F., Fowler, A., Di Francia, A., Panovska-Griffiths, J., et al.: Digital measurement of sars-cov-2 transmission risk from 7 million contacts. Nature 626(7997), 145–150 (2024)

[35] Eames, K.T., Tilston, N.L., Brooks-Pollock, E., Edmunds, W.J.: Measured dynamic social contact patterns explain the spread of h1n1v influenza. PLoS Computational Biology 8(3), 1002425 (2012)

[36] Wong, K.L., Gimma, A., Coletti, P., Faes, C., Beutels, P., Hens, N., Jaeger, V.K., Karch, A., Johnson, H., et al.: Social contact patterns during the covid-19 pandemic in 21 european countries–evidence from a two-year study. BMC Infectious Diseases 23(1), 268 (2023)

[37] Kummer, A.G., Zhang, J., Jiang, C., Litvinova, M., Ventura, P.C., Garcia, M.A., Vespignani, A., Wu, H., Yu, H., Ajelli, M.: Evaluating seasonal variations in human contact patterns and their impact on the transmission of respiratory infectious diseases. Influenza and Other Respiratory Viruses 18(5), 13301 (2024)

[28] Schuppert, A., Polotzek, K., Schmitt, J., Busse, R., Karschau, J., Karagiannidis, C.: Different spreading dynamics throughout Germany during the second wave of the COVID-19 pandemic: A time series study based on national surveillance data. The Lancet Regional Health - Europe 6, 100151 (2021) 10.1016/j.lanepe.2021.100151

[39] Pastor-Satorras, R., Castellano, C., Van Mieghem, P., Vespignani, A.: Epidemic processes in complex networks. Reviews of Modern Physics 87(3), 925–979 (2015) 10.1103/RevModPhys.87.925

[40] Keeling, M.J., Eames, K.T.D.: Networks and epidemic models. Journal of the Royal Society Interface 2(4), 295–307 (2005) 10.1098/rsif.2005.0051

[41] Cohen, R., Erez, K., ben-Avraham, D., Havlin, S.: Breakdown of the Internet under Intentional Attack. Physical Review Letters 86(16), 3682–3685 (2001) 10.1103/physrevlett.86.3682

[8] Kerr, C.C., Stuart, R.M., Mistry, D., Abeysuriya, R.G., Rosenfeld, K., Hart, G.R., Núñez, R.C., Cohen, J.A., Selvaraj, P., Hagedorn, B., George, L., Jastrzebski, M., Izzo, A.S., Fowler, G., Palmer, A., Delport, D., Scott, N., Kelly, S.L., Bennette, C.S., Wagner, B.G., Chang, S.T., Oron, A.P., Wenger, E.A., Panovska-Griffiths, J., Famulare, M., Klein, D.J.: Covasim: An agent-based model of COVID-19 dynamics and interventions. PLoS Computational Biology 17(7), 1009149 (2021) 10.1371/journal.pcbi.1009149

[43] Leng, T., White, C., Hilton, J., Kucharski, A., Pellis, L., Stage, H., Davies, N.G., Keeling, M.J., Flasche, S., et al.: The effectiveness of social bubbles as part of a covid-19 lockdown exit strategy, a modelling study. Wellcome Open Research 5, 213 (2021)

[44] Jarvis, C.I., Gimma, A., Wong, K.L.M., Van Zandvoort, J., Munday, J.D., Klepac, P., Funk, S., Edmunds, W.J.: Comix – individuals with high numbers of contacts (2021)

[45] Loedy, N., Coletti, P., Wambua, J., Hermans, L., Willem, L., Jarvis, C.I., Wong, K.L.M., Edmunds, W.J., Robert, A., Leclerc, Q.J., Gimma, A., Molenberghs, G., Beutels, P., Faes, C., Hens, N.: Longitudinal social contact data analysis: insights from 2 years of data collection in belgium during the COVID-19 pandemic. BMC Public Health 23(1) (2023) 10.1186/s12889-023-16193-7

[46] Backer, J.A., Vos, E.R., Hartog, G., Hagen, C.C., Melker, H.E., Klis, F.R., Wallinga, J.: Contact behaviour before, during and after the covid-19 pandemic in the netherlands: evidence from contact surveys, 2016 to 2017 and 2020 to 2023. Eurosurveillance 29(43), 2400143 (2024)

[47] Newman, M.: Power laws, pareto distributions and zipf’s law. Contemporary Physics 46(5), 323–351 (2005) 10.1080/00107510500052444

[48] Kenah, E., Robins, J.M.: Second look at the spread of epidemics on networks. Physical Review E 76(3), 036113 (2007)

[49] Cohen, R., Havlin, S., Ben-Avraham, D.: Efficient immunization strategies for computer networks and populations. Physical Review Letters 91(24), 247901 (2003)

[50] Sinclair, R.R., Allen, T., Barber, L., Bergman, M., Britt, T., Butler, A., Ford, M., Hammer, L., Kath, L., Probst, T., et al.: Occupational health science in the time of covid-19: Now more than ever. Occupational Health Science 4(1), 1–22 (2020)

[51] Kain, M.P., Childs, M.L., Becker, A.D., Mordecai, E.A.: Chopping the tail: how preventing superspreading can help to maintain covid-19 control. Epidemics 34, 100430 (2021)

[52] Meyers, L.A., Pourbohloul, B., Newman, M.E.J., Skowronski, D.M., Brunham, R.C.: Network theory and SARS: Predicting outbreak diversity. Journal of Theoretical Biology 232(1), 71–81 (2005) 10.1016/j.jtbi.2004.07.026

[53] Jin, Z., Zhang, J., Song, L.-P., Sun, G.-Q., Kan, J., Zhu, H.: Modelling and analysis of influenza A (H1N1) on networks. BMC Public Health 11(S1) (2011) 10.1186/1471-2458-11-s1-s9

[54] Karaivanov, A.: A social network model of COVID-19. PLoS ONE 15(10), 0240878 (2020) 10.1371/journal.pone.0240878

[55] Maheshwari, P., Albert, R.: Network model and analysis of the spread of COVID-19 with social distancing. Applied Network Science 5(1) (2020) 10.1007/s41109-020-00344-5

[56] Cui, Y., Ni, S., Shen, S.: A network-based model to explore the role of testing in the epidemiological control of the COVID-19 pandemic. BMC Infectious Diseases 21(1) (2021) 10.1186/s12879-020-05750-9

[57] Zonta, F., Levitt, M.: Virus spread on a scale-free network reproduces the Gompertz growth observed in isolated COVID-19 outbreaks. Advances in Biological Regulation 86, 100915 (2022) 10.1016/j.jbior.2022.100915

[58] Cheng, S., Pain, C.C., Guo, Y.-K., Arcucci, R.: Real-time updating of dynamic social networks for COVID-19 vaccination strategies. Journal of Ambient Intelligence and Humanized Computing 15(3), 1981–1994 (2024) 10.1007/s12652-023-04589-7

[59] Schnitzler, L., Janssen, L.M., Evers, S.M., Jackson, L.J., Paulus, A.T., Roberts, T.E., Pokhilenko, I.: The broader societal impacts of covid-19 and the growing importance of capturing these in health economic analyses. International Journal of Technology Assessment in Health Care 37(1), 43 (2021)

[60] Cauchemez, S., Bhattarai, A., Marchbanks, T.L., Fagan, R.P., Ostroff, S., Ferguson, N.M., Swerdlow, D., Group, P.H.W., Sodha, S.V., Moll, M.E., et al.: Role of social networks in shaping disease transmission during a community outbreak of 2009 h1n1 pandemic influenza. Proceedings of the National Academy of Sciences 108(7), 2825–2830 (2011)

[61] Knabl, L., Mitra, T., Kimpel, J., Rössler, A., Volland, A., Walser, A., Ulmer, H., Pipperger, L., Binder, S.C., Riepler, L., Bates, K., Bandyopadhyay, A., Schips, M., Ranjan, M., Falkensammer, B., Borena, W., Meyer-Hermann, M., Laer, D.: High sars-cov-2 seroprevalence in children and adults in the austrian ski resort of ischgl. Communications Medicine 1(1), 4 (2021) 10.1038/s43856-021-00007-1

[62] Günther, T., Czech-Sioli, M., Indenbirken, D., Robitaille, A., Tenhaken, P., Exner, M., Ottinger, M., Fischer, N., Grundhoff, A., Brinkmann, M.M., et al.: Sars-cov-2 outbreak investigation in a german meat processing plant. EMBO Molecular Medicine 12(12), 13296 (2020) 10.15252/emmm.202013296

[63] Rhodes, S., Wilkinson, J., Pearce, N., Mueller, W., Cherrie, M., Stocking, K., Gittins, M., Katikireddi, S.V., Tongeren, M.V.: Occupational differences in SARS-CoV-2 infection: analysis of the UK ONS COVID-19 infection survey. J. Epidemiol. Community Health 76(10), 841–846 (2022) 10.1136/jech-2022-219101

[64] Eekhout, I., Tongeren, M., Pearce, N., Oude Hengel, K.M.: The impact of occupational exposures on infection rates during the covid-19 pandemic: a test-negative design study with register data of 207 034 dutch workers. Scandinavian Journal of Work, Environment & Health (4), 259–270 (2023) 10.5271/sjweh.4086

[65] Chang, S., Pierson, E., Koh, P.W., Gerardin, J., Redbird, B., Grusky, D., Leskovec, J.: Mobility network models of covid-19 explain inequities and inform reopening. Nature 589(7840), 82–87 (2021) 10.1038/s41586-020-2923-3

[66] Fan, C., Lee, R., Yang, Y., Mostafavi, A.: Fine-grained data reveal segregated mobility networks and opportunities for local containment of covid-19. Scientific Reports 11, 16895 (2021) 10.1038/s41598-021-95894-8

[67] Santana, C., Botta, F., Barbosa, H., Privitera, F., Menezes, R., DiClemente, R.: Covid-19 is linked to changes in the time–space dimension of human mobility. Nature Human Behaviour 7(10), 1729–1739 (2023) 10.1038/s41562-023-01660-3

[68] Riley, S., Fraser, C., Donnelly, C.A., Ghani, A.C., Abu-Raddad, L.J., Hedley, A.J., Leung, G.M., Ho, L.-M., Lam, T.-H., Thach, T.Q., Chau, P., Chan, K.-P., Lo, S.-V., Leung, P.-Y., Tsang, T., Ho, W., Lee, K.-H., Lau, E.M.C., Ferguson, N.M., Anderson, R.M.: Transmission Dynamics of the Etiological Agent of SARS in Hong Kong: Impact of Public Health Interventions. Science 300(5627), 1961–1966 (2003) 10.1126/science.1086478

[69] Elliott, M.R., Valliant, R.: Inference for Nonprobability Samples. Statistical Science 32(2) (2017) 10.1214/16-STS598

[70] Bethlehem, J.: Selection Bias in Web Surveys. International Statistical Review 78(2), 161–188 (2010) 10.1111/j.1751-5823.2010.00112.x

[65] Dan, S., Ling, Z., Chen, Y., Tegegne, J., Jaeger, V.K., Karch, A., Mishra, S., Ratmann, O., The Machine Learning &Global Health network, Bhatt, S., Flaxman, S., Semenova, E., Miscouridou, X., Unwin, J., Sejdinovic, D., Faria, N., Duchene, D.A., Kraemer, M., Vollmer, S.: Addressing survey fatigue bias in longitudinal social contact studies to improve pandemic preparedness. Scientific Reports 15(1), 17935 (2025) 10.1038/s41598-025-02235-0

[72] Balcan, D., Colizza, V., Gonçalves, B., Hu, H., Ramasco, J.J., Vespignani, A.: Modeling the spatial spread of infectious diseases: the global epidemic and mobility computational model. BMC Medicine 7, 45 (2009) 10.1186/1741-7015-7-45

[73] Ipsos iSay. https://www.ipsos.com

[74] Gimma, A., Munday, J.D., Wong, K.L.M., Coletti, P., Zandvoort, K., Prem, K., Klepac, P., Rubin, G.J., Funk, S., Edmunds, W.J., and, C.I.J.: Changes in social contacts in england during the COVID-19 pandemic between march 2020 and march 2021 as measured by the CoMix survey: A repeated crosssectional study. PLoS Medicine 19(3), 1003907 (2022) 10.1371/journal.pmed.1003907

[75] Phuong, H.T., Bartz, A., Jarynowski, A.K., Lange, B., Jarvis, C.I., Rübsamen, N., Mikolajczyk, R.T., Scholz, S., Berger, T., Heinsohn, T., Belik, V., Karch, A., Jaeger, V.K.: Changes in social contact patterns in Germany during the SARS-CoV-2 pandemic – an analysis based on the COVIMOD study. BMC Infectious Diseases 25(1), 588 (2025) 10.1186/s12879-025-10917-3

[32] Gelman, A.: Poststratification into many categories using hierarchical logistic regression. Survey Methodology 23, 127 (1997)

[24] Stan Development Team: Prior Choice Recommendations. Stan Wiki (2025). https://github.com/stan-dev/stan/wiki/Prior-Choice-Recommendations

[5] Statistisches Bundesamt (Destatis): GENESIS-Online Datenbank. https://www-genesis.destatis.de/datenbank/online

[33] Ling, Z., Dan, S.: Bayesian Shrinkage Priors Subject to Linear Constraints. arXiv (2025). 10.48550/arXiv.2504.09052

[34] Mistry, D., Litvinova, M., Pastore Y Piontti, A., Chinazzi, M., Fumanelli, L., Gomes, M.F.C., Haque, S.A., Liu, Q.-H., Mu, K., Xiong, X., Halloran, M.E., Longini, I.M., Merler, S., Ajelli, M., Vespignani, A.: Inferring high-resolution human mixing patterns for disease modeling. Nature Communications 12(1) (2021) 10.1038/s41467-020-20544-y

[25] Vehtari, A., Gelman, A., Gabry, J.: Practical bayesian model evaluation using leave-one-out crossvalidation and WAIC. Statistics and Computing 27(5), 1413–1432 (2017) 10.1007/s11222-016-9696-4

[66] Piironen, J., Vehtari, A.: Sparsity information and regularization in the horseshoe and other shrinkage priors. Electronic Journal of Statistics 11(2) (2017) 10.1214/17-EJS1337SI

## References

[1] Ensheng Dong, Hongru Du, and Lauren Gardner. An interactive web-based dashboard to track covid-19 in real time. The Lancet Infectious Diseases, 20(5):533–534, 2020. ISSN 1473-3099. doi: 10.1016/S1473-3099(20)30120-1. URL https://doi.org/10.1016/s1473-3099(20)30120-1.

[2] OpenStreetMap contributors. Planet dump retrieved from https://planet.osm.org. https://www.openstreetmap.org, 2017.

[3] Opendatasoft. Kreise–germany, 2020. URL https://public.opendatasoft.com/explore/dataset/georef-germany-kreis/.

[4] NUTS–Nomenclature of Territorial Units for Statistics. https://ec.europa.eu/eurostat/web/nuts/overview, 2024.

[5] Statistisches Bundesamt (Destatis). Genesis-online datenbank, 2024. URL https://www-genesis.destatis.de/datenbank/online.

[6] Matthew D. Hoffman and Andrew Gelman. The No-U-Turn sampler: Adaptively setting path lengths in Hamiltonian Monte Carlo, 2014. URL https://arxiv.org/abs/1111.4246.

[7] Bob Carpenter, Andrew Gelman, Matthew D. Hoffman, Daniel Lee, Ben Goodrich, Michael Betancourt, Marcus Brubaker, Jiqiang Guo, Peter Li, and Allen Riddell. Stan: A probabilistic programming language. Journal of Statistical Software, 76(1), 2017. doi: 10.18637/jss.v076.i01. URL https://doi.org/10.18637/jss.v076.i01.

[8] Cliff C. Kerr, Robyn M. Stuart, Dina Mistry, Romesh G. Abeysuriya, Katherine Rosenfeld, Gregory R. Hart, Rafael C. Núñez, Jamie A. Cohen, Prashanth Selvaraj, Brittany Hagedorn, Lauren George, Michal Jastrzebski, Amanda S. Izzo, Greer Fowler, Anna Palmer, Dominic Delport, Nick Scott, Sherrie L. Kelly, Caroline S. Bennette, Bradley G. Wagner, Stewart T. Chang, Assaf P. Oron, Edward A. Wenger, Jasmina Panovska-Griffiths, Michael Famulare, and Daniel J. Klein. Covasim: An agent-based model of COVID-19 dynamics and interventions. PLOS Computational Biology, 17(7):e1009149, July 2021. ISSN 1553-7358. doi: 10.1371/journal.pcbi.1009149.

[9] C. D. Kemp and A. W. Kemp. Generalized Hypergeometric Distributions. 18(2):202–211. ISSN 2517-6161. doi: 10.1111/j.2517-6161.1956.tb00224.x.

[10] J. O. Irwin. The generalized waring distribution applied to accident theory. Journal of the Royal Statistical Society Series A, 131(2):205, 1968. doi: 10.2307/2343842. URL https://doi.org/10.2307/2343842.

[11] J. O. Irwin. The generalized waring distribution. part i. Journal of the Royal Statistical Society Series A, 138(1):18, 1975. doi: 10.2307/2345247. URL https://doi.org/10.2307/2345247.

[12] J. O. Irwin. The generalized waring distribution. part II. Journal of the Royal Statistical Society Series A, 138(2):204, 1975. doi: 10.2307/2984648. URL https://doi.org/10.2307/2984648.

[13] J. O. Irwin. The generalized waring distribution. part III. Journal of the Royal Statistical Society Series A, 138(3):374, 1975. doi: 10.2307/2344582. URL https://doi.org/10.2307/2344582.

[14] Evdokia Xekalaki. Chance mechanisms for the univariate generalized waring distribution and related characterizations. pages 157–171, 1981. doi: 10.1007/978-94-009-8549-0_12. URL https://doi.org/10.1007/978-94-009-8549-0_12.

[15] Evdokia Xekalaki. Infinite divisibility, completeness and regression properties of the univariate generalized waring distribution. Annals of the Institute of Statistical Mathematics, 35(2):279–289, December 1983. doi: 10.1007/bf02480983. URL https://doi.org/10.1007/bf02480983.

[16] Evdokia Xekalaki. The univariate generalized waring distribution in relation to accident theory: Proneness, spells or contagion? Biometrics, 39(4):887, December 1983. doi: 10.2307/2531324. URL https://doi.org/10.2307/2531324.

[17] Evdokia Xekalaki. A property of the yule distribution and its applications. Communications in Statistics Theory and Methods, 12(10):1181–1189, January 1983. doi: 10.1080/03610928308828523. URL https://doi.org/10.1080/03610928308828523.

[18] Evdokia Xekalaki. Hazard functions and life distributions in discrete time. Communications in Statistics Theory and Methods, 12(21):2503–2509, January 1983. doi: 10.1080/03610928308828617. URL https://doi.org/10.1080/03610928308828617.

[19] James Holland Jones and Mark S. Handcock. Sexual contacts and epidemic thresholds. 423(6940): 605–606. ISSN 0028-0836, 1476-4687. doi: 10.1038/423605a.

[20] J. H. Jones and M. S. Handcock. An assessment of preferential attachment as a mechanism for human sexual network formation. Proceedings of the Royal Society of London. Series B: Biological Sciences, 270(1520):1123–1128, June 2003. ISSN 0962-8452, 1471-2954. doi: 10.1098/rspb.2003.2369.

[21] Mark S. Handcock and James Holland Jones. Likelihood-based inference for stochastic models of sexual network formation. Theoretical Population Biology, 65(4):413–422, June 2004. ISSN 0040-5809. doi: 10.1016/j.tpb.2003.09.006.

[22] Andrew J. Leigh Brown, Samantha J. Lycett, Lucy Weinert, Gareth J. Hughes, Esther Fearnhill, David T. Dunn, and on behalf of the UK HIV Drug Resistance Collaboration. Transmission Network Parameters Estimated From HIV Sequences for a Nationwide Epidemic. The Journal of Infectious Diseases, 204(9):1463–1469, November 2011. ISSN 0022-1899. doi: 10.1093/infdis/jir550.

[23] Gabriel Riutort-Mayol, Paul-Christian Bürkner, Michael R. Andersen, Arno Solin, and Aki Vehtari. Practical Hilbert space approximate Bayesian Gaussian processes for probabilistic programming. 33(1): 17. ISSN 0960-3174, 1573-1375. doi: 10.1007/s11222-022-10167-2.

[24] Stan Development Team. Prior choice recommendations. Stan Wiki, 2025. URL https://github.com/stan-dev/stan/wiki/Prior-Choice-Recommendations.

[25] Aki Vehtari, Andrew Gelman, and Jonah Gabry. Practical bayesian model evaluation using leave-one-out cross-validation and WAIC. Statistics and Computing, 27(5):1413–1432, August 2017. doi: 10.1007/s11222-016-9696-4. URL https://doi.org/10.1007/s11222-016-9696-4.

[26] Aki Vehtari, Daniel Simpson, Andrew Gelman, Yuling Yao, and Jonah Gabry. Pareto Smoothed Importance Sampling, March 2024.

[27] Fedelis Mutiso, Hong Li, John L Pearce, Sara E Benjamin-Neelon, Noel T Mueller, and Brian Neelon. Bayesian kernel machine regression for count data: Modelling the association between social vulnerability and COVID-19 deaths in South Carolina. Journal of the Royal Statistical Society Series C, 73(1): 257–274, January 2024. ISSN 0035-9254, 1467-9876. doi: 10.1093/jrsssc/qlad094.

[28] Andreas Schuppert, Katja Polotzek, Jochen Schmitt, Reinhard Busse, Jens Karschau, and Christian Karagiannidis. Different spreading dynamics throughout Germany during the second wave of the COVID-19 pandemic: A time series study based on national surveillance data. 6:100151. ISSN 26667762. doi: 10.1016/j.lanepe.2021.100151.

[29] Statistisches Bundesamt (Destatis). Population pyramid: Age structure of the population in germany, 2021. URL https://service.destatis.de/bevoelkerungspyramide/index.html#!y=2021.

[30] D. Holt and T. M. F. Smith. Post Stratification. Journal of the Royal Statistical Society Series A, 142 (1):33, 1979. ISSN 00359238. doi: 10.2307/2344652.

[31] R. J. A. Little. Post-Stratification: A Modeler’s Perspective. 88(423):1001–1012. ISSN 0162-1459, 1537-274X. doi: 10.1080/01621459.1993.10476368.

[32] Andrew Gelman. Poststratification into many categories using hierarchical logistic regression. Survey methodology, 23:127, 1997.

[33] Zhi Ling and Shozen Dan. Bayesian shrinkage priors subject to linear constraints, April 2025. URL https://arxiv.org/abs/2504.09052.

[34] Dina Mistry, Maria Litvinova, Ana Pastore Y Piontti, Matteo Chinazzi, Laura Fumanelli, Marcelo F. C. Gomes, Syed A. Haque, Quan-Hui Liu, Kunpeng Mu, Xinyue Xiong, M. Elizabeth Halloran, Ira M. Longini, Stefano Merler, Marco Ajelli, and Alessandro Vespignani. Inferring high-resolution human mixing patterns for disease modeling. Nature Communications, 12(1), January 2021. ISSN 2041-1723. doi: 10.1038/s41467-020-20544-y.

[35] M. E. J. Newman. Spread of epidemic disease on networks. Physical Review E, 66(1):016128, July 2002. ISSN 1063-651X, 1095-3787. doi: 10.1103/PhysRevE.66.016128.

[36] Hualei Xin, Zhe Wang, Shuang Feng, Zhou Sun, Lele Yu, Benjamin J Cowling, Qingxin Kong, and Peng Wu. Transmission dynamics of SARS-CoV-2 Omicron variant infections in Hangzhou, Zhejiang, China, January-February 2022. International Journal of Infectious Diseases, 126:132–135, January 2023. ISSN 12019712. doi: 10.1016/j.ijid.2022.10.033.

[37] Jiasheng Shao, Rong Fan, Jianrong Hu, Tiejun Zhang, Catherine Lee, Xuyuan Huang, Fei Wang, Haiying Liang, Ye Jin, Ying Jiang, Yanhua Gu, and Gang Huang. Clinical Progression and Outcome of Hospitalized Patients Infected with SARS-CoV-2 Omicron Variant in Shanghai, China. Vaccines, 10 (9):1409, August 2022. ISSN 2076-393X. doi: 10.3390/vaccines10091409.

[38] UK Health Security Agency. Covid-19 infectious period: a rapid evidence review (update 1). Rapid evidence review, UK Health Security Agency, January 2024. URL https://assets.publishing.service.gov.uk/media/658580c723b70a0013234e62/COVID-19-infectious-period-a-rapid-evidence-review-update-1.pdf. Accessed: 2025-12-07.

[39] Jacqui Wise. Covid-19: Symptomatic infection with omicron variant is milder and shorter than with delta, study reports. BMJ, page o922, April 2022. ISSN 1756-1833. doi: 10.1136/bmj.o922.

[40] Jun Cai, Xiaowei Deng, Juan Yang, Kaiyuan Sun, Hengcong Liu, Zhiyuan Chen, Cheng Peng, Xinhua Chen, Qianhui Wu, Junyi Zou, Ruijia Sun, Wen Zheng, Zeyao Zhao, Wanying Lu, Yuxia Liang, Xiaoyu Zhou, Marco Ajelli, and Hongjie Yu. Modeling transmission of SARS-CoV-2 Omicron in China. Nature Medicine, 28(7):1468–1475, July 2022. ISSN 1078-8956, 1546-170X. doi: 10.1038/s41591-022-01855-7.

[41] Keerthana Sankar, Neil Modi, Alexander Polyak, Michael P Directo, Lily R Johnson, Norling Kho, Sharon K Isonaka, Isabel Pedraza, Peter Chen, and Matthew E Modes. Comparison of clinical characteristics and outcomes of critically ill adults with SARS-CoV-2 infection during Delta and Omicron variant predominance periods: A single-hospital retrospective cohort study. BMJ Open Respiratory Research, 10(1):e001274, February 2023. ISSN 2052-4439. doi: 10.1136/bmjresp-2022-001274.

[42] World Health Organization. Influenza (seasonal), 2023. URL https://www.who.int/news-room/fact-sheets/detail/influenza-(seasonal). Accessed: 2025-12-07.

[43] Centers for Disease Control and Prevention. Influenza (seasonal, zoonotic, and pandemic) – cdc yellow book 2024, 2023. URL https://www.cdc.gov/yellow-book/hcp/travel-associated-infections-diseases/influenza.html. Accessed: 2025-12-07.

[44] A. Campbell, R. Rodin, R. Kropp, Y. Mao, Z. Hong, J. Vachon, J. Spika, and L. Pelletier. Risk of severe outcomes among patients admitted to hospital with pandemic (H1N1) influenza. Canadian Medical Association Journal, 182(4):349–355, March 2010. ISSN 0820-3946, 1488-2329. doi: 10.1503/cmaj.091823.

[45] Timothée Abaziou, Clément Delmas, Fanny Vardon Bounes, Fabien Bignon, Laure Crognier, Thierry Seguin, Béatrice Riu-Poulenc, Stéphanie Ruiz, Antoine Rouget, Pierre Cougot, Bernard Georges, Jean-Marie Conil, and Vincent Minville. Outcome of Critically Ill Patients With Influenza Infection: A Retrospective Study. Infectious Diseases: Research and Treatment, 13:1178633720904081, January 2020. ISSN 1178-6337, 1178-6337. doi: 10.1177/1178633720904081.

[46] Muriel Fartoukh, Guillaume Voiriot, Laurent Guérin, Jean Damien Ricard, Alain Combes, Morgane Faure, Sarah Benghanem, Etienne De Montmollin, Yacine Tandjaoui-Lambiotte, Antoine Vieillard-Baron, Eric Maury, Jean-Luc Diehl, Keyvan Razazi, Virginie Lemiale, Pierre Trouiller, Benjamin Planquette, Laurent Savale, Nicholas Heming, Jonathan Marey, AP-HP Clinical Data Warehouse, Fabrice Carrat, Nathanael Lapidus, The EPIcuFLU APHP Group, Michel Djibré, Jean Louis Teboul, Jonathan Messika, Alexandre Demoule, Jean Paul Mira, Jean-François Timsit, Yves Cohen, Bernard Page, Armand Mekontso Dessap, Elie Azoulay, Olivier Sanchez, Marc Humbert, Djillali Annane, and Nicolas Roche. Seasonal burden of severe influenza virus infection in the critically ill patients, using the Assistance Publique-Hôpitaux de Paris clinical data warehouse: A pilot study. Annals of Intensive Care, 11(1):117, December 2021. ISSN 2110-5820. doi: 10.1186/s13613-021-00884-8.

[47] Fabrice Carrat, Elisabeta Vergu, Neil M. Ferguson, Magali Lemaitre, Simon Cauchemez, Steve Leach, and Alain-Jacques Valleron. Time Lines of Infection and Disease in Human Influenza: A Review of Volunteer Challenge Studies. American Journal of Epidemiology, 167(7):775–785, April 2008. ISSN 1476-6256, 0002-9262. doi: 10.1093/aje/kwm375.

[48] Centers for Disease Control and Prevention. Clinical signs and symptoms of influenza, 2024. URL https://www.cdc.gov/flu/hcp/clinical-signs/index.html. Accessed: 2025-12-07.

[49] Alison Han, Lindsay M Czajkowski, Amanda Donaldson, Holly Ann Baus, Susan M Reed, Rani S Athota, Tyler Bristol, Luz Angela Rosas, Adriana Cervantes-Medina, Jeffery K Taubenberger, and Matthew J Memoli. A Dose-finding Study of a Wild-type Influenza A(H3N2) Virus in a Healthy Volunteer Human Challenge Model. Clinical Infectious Diseases, 69(12):2082–2090, November 2019. ISSN 1058-4838, 1537-6591. doi: 10.1093/cid/ciz141.

[50] Mathilde Pivette, Nathalie Nicolay, Virginie De Lauzun, and Bruno Hubert. Characteristics of hospitalizations with an influenza diagnosis, France, 2012-2013 to 2016-2017 influenza seasons. Influenza and Other Respiratory Viruses, 14(3):340–348, May 2020. ISSN 1750-2640, 1750-2659. doi: 10.1111/irv.12719.

[51] Yousef Alimohamadi, Maryam Taghdir, and Mojtaba Sepandi. Estimate of the Basic Reproduction Number for COVID-19: A Systematic Review and Meta-analysis. Journal of Preventive Medicine and Public Health, 53(3):151–157, May 2020. ISSN 1975-8375, 2233-4521. doi: 10.3961/jpmph.20.076.

[52] Nicholas G. Davies, Petra Klepac, Yang Liu, Kiesha Prem, Mark Jit, CMMID COVID-19 working group, Carl A. B. Pearson, Billy J. Quilty, Adam J. Kucharski, Hamish Gibbs, Samuel Clifford, Amy Gimma, Kevin Van Zandvoort, James D. Munday, Charlie Diamond, W. John Edmunds, Rein M. G. J. Houben, Joel Hellewell, Timothy W. Russell, Sam Abbott, Sebastian Funk, Nikos I. Bosse, Yueqian Fiona Sun, Stefan Flasche, Alicia Rosello, Christopher I. Jarvis, and Rosalind M. Eggo. Age-dependent effects in the transmission and control of COVID-19 epidemics. Nature Medicine, 26(8):1205–1211, August 2020. ISSN 1078-8956, 1546-170X. doi: 10.1038/s41591-020-0962-9.

[53] Neil Ferguson, daniel Laydon, gemma Nedjati Gilani, Natsuko Imai, kylie Ainslie, marc Baguelin, sangeeta Bhatia, adhiratha Boonyasiri, zulma Cucunuba Perez, gina Cuomo-Dannenburg, amy Dighe, ilaria Dorigatti, han Fu, katy Gaythorpe, will Green, arran Hamlet, wes Hinsley, lucy Okell, Sabine van Elsland, hayley Thompson, robert Verity, erik Volz, haowei Wang, Yuanrong Wang, patrick Walker, Caroline Walters, peter Winskill, charles Whittaker, christl Donnelly, steven Riley, and azra Ghani. Report 9: Impact of non-pharmaceutical interventions (NPIs) to reduce COVID19 mortality and healthcare demand. March 2020. doi: 10.25561/77482.

[54] Megan O’Driscoll, Gabriel Ribeiro Dos Santos, Lin Wang, Derek A. T. Cummings, Andrew S. Azman, Juliette Paireau, Arnaud Fontanet, Simon Cauchemez, and Henrik Salje. Age-specific mortality and immunity patterns of SARS-CoV-2. Nature, 590(7844):140–145, February 2021. ISSN 0028-0836, 1476-4687. doi: 10.1038/s41586-020-2918-0.

[55] Daihai He, Shi Zhao, Qianying Lin, Zian Zhuang, Peihua Cao, Maggie H. Wang, and Lin Yang. The relative transmissibility of asymptomatic COVID-19 infections among close contacts. International Journal of Infectious Diseases, 94:145–147, May 2020. ISSN 12019712. doi: 10.1016/j.ijid.2020.04.034.

[56] Ying Liu and Joacim Rocklöv. The effective reproductive number of the Omicron variant of SARS-CoV-2 is several times relative to Delta. Journal of Travel Medicine, 29(3):taac037, May 2022. ISSN 1195-1982, 1708-8305. doi: 10.1093/jtm/taac037.

[57] Weien Yu, Yifei Guo, Shenyan Zhang, Yide Kong, Zhongliang Shen, and Jiming Zhang. Proportion of asymptomatic infection and nonsevere disease caused by SARS-CoV-2 Omicron variant: A systematic review and analysis. Journal of Medical Virology, 94(12):5790–5801, December 2022. ISSN 0146-6615, 1096-9071. doi: 10.1002/jmv.28066.

[58] Chia Siang Kow, Dinesh Sangarran Ramachandram, and Syed Shahzad Hasan. The risk of mortality and severe illness in patients infected with the omicron variant relative to delta variant of SARS-CoV-2: A systematic review and meta-analysis. Irish Journal of Medical Science (1971-), 192(6):2897–2904, December 2023. ISSN 0021-1265, 1863-4362. doi: 10.1007/s11845-022-03266-6.

[59] Ji-Eun Park and Yeonhee Ryu. Transmissibility and severity of influenza virus by subtype. Infection, Genetics and Evolution, 65:288–292, November 2018. ISSN 15671348. doi: 10.1016/j.meegid.2018.08.007.

[60] Louis Yat Hin Chan, Sinead E. Morris, Melissa S. Stockwell, Natalie M. Bowman, Edwin Asturias, Suchitra Rao, Karen Lutrick, Katherine D. Ellingson, Huong Q. Nguyen, Yvonne Maldonado, Son H. McLaren, Ellen Sano, Jessica E. Biddle, Sarah E. Smith-Jeffcoat, Matthew Biggerstaff, Melissa A. Rolfes, H. Keipp Talbot, Carlos G. Grijalva, Rebecca K. Borchering, and Alexandra M. Mellis. Estimating the generation time for influenza transmission using household data in the United States. Epidemics, 50: 100815, March 2025. ISSN 17554365. doi: 10.1016/j.epidem.2025.100815.

[61] Scott A. McDonald, Anne C. Teirlinck, Mariette Hooiveld, Liselotte Van Asten, Adam Meijer, Marit De Lange, Arianne B. Van Gageldonk-Lafeber, and Jacco Wallinga. Inference of age-dependent case-fatality ratios for seasonal influenza virus subtypes A(H3N2) and A(H1N1)pdm09 and B lineages using data from the Netherlands. Influenza and Other Respiratory Viruses, 17(6):e13146, June 2023. ISSN 1750-2640, 1750-2659. doi: 10.1111/irv.13146.

[62] Tim K. Tsang, Can Wang, Vicky J. Fang, Ranawaka A. P. M. Perera, Hau Chi So, Dennis K. M. Ip, Gabriel M. Leung, J. S. Malik Peiris, Simon Cauchemez, and Benjamin J. Cowling. Reconstructing household transmission dynamics to estimate the infectiousness of asymptomatic influenza virus infections. Proceedings of the National Academy of Sciences, 120(33):e2304750120, August 2023. ISSN 0027-8424, 1091-6490. doi: 10.1073/pnas.2304750120.

[63] Our World in Data. Daily covid-19 tests per 1,000 people, 2022. URL https://archive.ourworldindata.org/20250909-093708/grapher/daily-tests-per-thousand-people-smoothed-7-day.html.

[64] Nicole Ngai Yung Tsang, Hau Chi So, Ka Yan Ng, Benjamin J Cowling, Gabriel M Leung, and Dennis Kai Ming Ip. Diagnostic performance of different sampling approaches for SARS-CoV-2 RT-PCR testing: A systematic review and meta-analysis. The Lancet Infectious Diseases, 21(9):1233–1245, September 2021. ISSN 14733099. doi: 10.1016/S1473-3099(21)00146-8.

[65] Shozen Dan, Zhi Ling, Yu Chen, Joshua Tegegne, Veronika K. Jaeger, André Karch, Swapnil Mishra, Oliver Ratmann, The Machine Learning & Global Health network, Samir Bhatt, Seth Flaxman, Elizaveta Semenova, Xenia Miscouridou, Juliette Unwin, Dino Sejdinovic, Nuno Faria, David A. Duchene, Moritz Kraemer, and Sebastian Vollmer. Addressing survey fatigue bias in longitudinal social contact studies to improve pandemic preparedness. Scientific Reports, 15(1):17935, May 2025. ISSN 2045-2322. doi: 10.1038/s41598-025-02235-0.

[66] Juho Piironen and Aki Vehtari. Sparsity information and regularization in the horseshoe and other shrinkage priors. 11(2). ISSN 1935-7524. doi: 10.1214/17-EJS1337SI.

